# Human systemic and mucosal immune responses support further exploration of a *Klebsiella pneumoniae* protein-based vaccine

**DOI:** 10.64898/2026.03.26.26349300

**Authors:** Joseph J Campo, Oliver Pearse, Allan M Zuza, Amit Oberai, Patricia Siyabu, Edith Tewesa, Luis Gadama, Samantha Lissauer, David Lissauer, Andy A Teng, Jozelyn V Pablo, Joshua M Edgar, Adam D Shandling, Kondwani Kawaza, Nicholas A Feasey, Eva Heinz

**Affiliations:** Antigen Discovery Incorporated, Irvine, California, USA; Malawi-Liverpool-Wellcome Programme, Kamuzu University of Health Sciences, Blantyre, Malawi; Department of Clinical Sciences, Liverpool School of Tropical Medicine, Liverpool, UK; Queen Elizabeth Central Hospital, Blantyre, Malawi; Kamuzu University of Health Sciences, Blantyre, Malawi; Institute Infection, Veterinary and Ecological Sciences, University of Liverpool, UK; Alder Hey Children’s Hospital, Liverpool, UK; Institute of Life Course & Medical Sciences, University of Liverpool, Liverpool, UK; School of Medicine, University of St. Andrews, St. Andrews, UK; Department of Vector Biology, Liverpool School of Tropical Medicine, Liverpool, UK; Strathclyde Institute of Pharmacy & Biomedical Sciences, University of Strathclyde, Glasgow, UK; Parasites and Microbes Program, Wellcome Sanger Institute, Hinxton, UK

**Keywords:** maternal vaccine, sub-Saharan Africa, neonatal sepsis

## Abstract

Neonatal sepsis caused by *Klebsiella pneumoniae* is a major cause of under-five mortality in sub-Saharan Africa, and the rapid increase of infections caused by bacteria resistant to most or all available antimicrobials severely limits treatment options. An effective, maternally-administered vaccine could make a substantial reduction in neonatal sepsis and associated negative outcomes, as well as reduce the overall need for antimicrobials, a key driver of antimicrobial resistance. This study explored the potential for a maternally administered protein-based vaccine to provide neonatal protection via antibodies transferred transplacentally and through breastfeeding. A case-control study of mother and baby dyads was designed with 20 neonates developing *K. pneumoniae* sepsis and 80 uninfected control neonates to analyse breastmilk IgA, cord blood IgG and maternal serum IgA and IgG antibodies on a protein microarray with 161 selected *K. pneumoniae* proteins representing 152 unique genes. This analysis identified a set of proteins eliciting antibody responses, some associated with lack of *K. pneumoniae* sepsis, that indicate the presence of potentially protective antibodies. This is an essential first step in exploring surface protein accessibility, despite the large capsule. We highlight fimbrial structures, conjugative pili, and small lipoproteins associated with large outer membrane complexes as potential protein vaccine targets.

**Graphical abstract:** 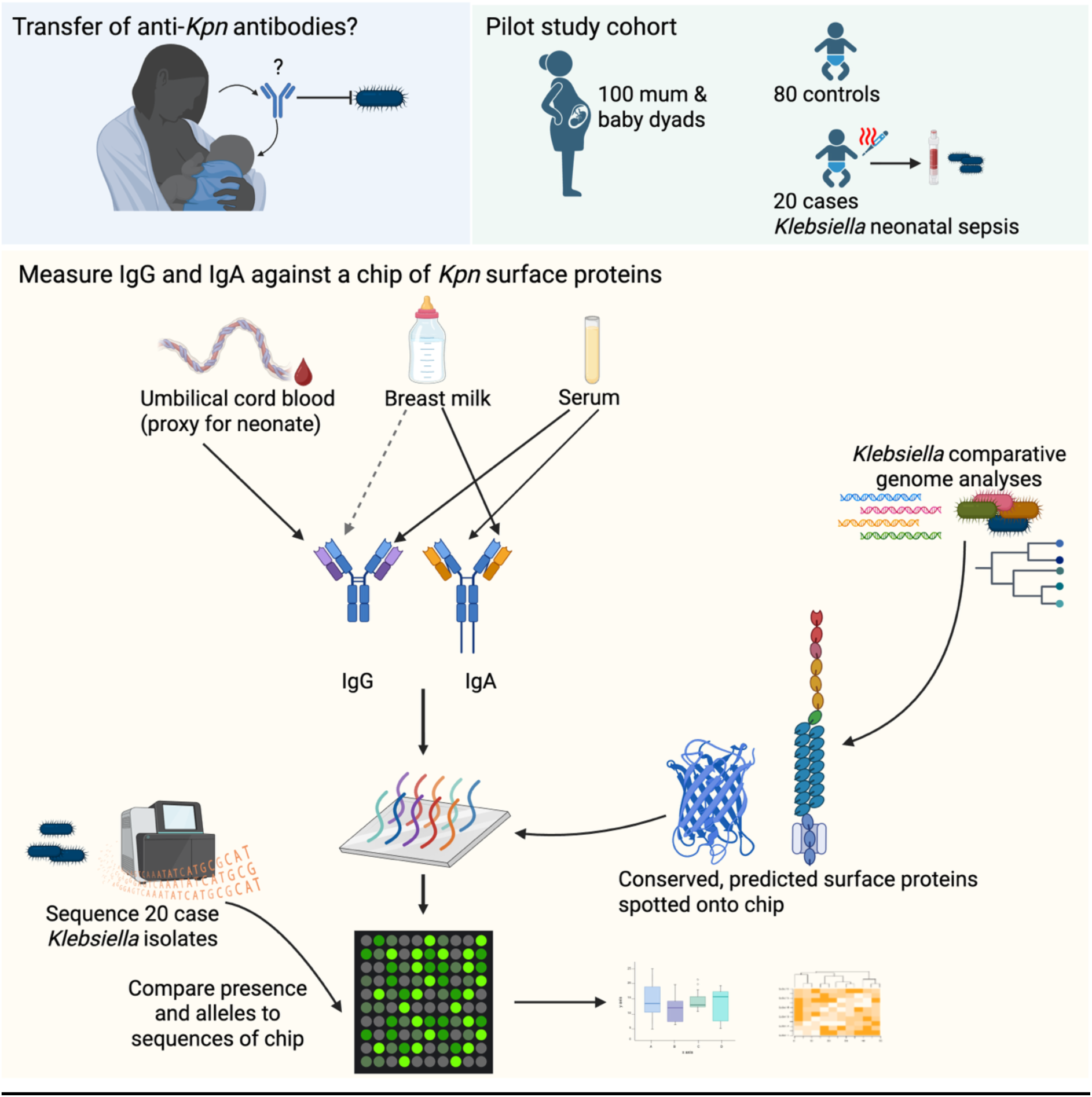

## Introduction

*Klebsiella pneumoniae* (*Kpn*) is the second most common cause of bacterial infection in neonates globally. Antimicrobial resistance (AMR) is highly prevalent in *Kpn*, causing serious difficulties in treatment in locations with limited access to World Health Organisation antibiotics designated Watch and Reserve. A specific issue is neonatal infections in low- and middle-income countries (LMICs), where neonatal sepsis is the leading cause of under-5 mortality(1, 2). This challenge is exacerbated by a rapid increase in prevalence of AMR amongst *Kpn*, particularly resistance to third-generation cephalosporins (3GC), often the last-line therapeutic option, especially in sub-Saharan Africa where prevalence of 3GC-resistance (3GC-R) has increased from 12% to >90% in some settings in the last decade(3).

Given the morbidity and mortality associated with neonatal sepsis even when caused by readily treatable bacteria, combined with the lack of access to potentially effective drugs and the strain on healthcare caused by neonatal sepsis, a vaccine is a highly attractive strategy to improve early life outcomes and lessen the burden on healthcare systems that are often operating beyond capacity. A maternally administered *Kpn* vaccine could be implemented in routine antenatal clinics for expecting mothers.

A key challenge in designing a vaccine against opportunistic pathogens like *Kpn*, which are not host-restricted but occur widely in the environment and can be a part of a healthy individuals’ microbiome, is their diversity. An attractive vaccine target for gram-negative bacteria is the surface polysaccharide layer, which is often highly immunogenic and has been successfully used for vaccines against major childhood pathogens like *Haemophilus influenzae* and *Streptococcus pneumonia*e. However, the *Kpn* capsule locus is highly diverse and the most recombinogenic region of the genome(4), currently with >180 K-loci identified. A recent large-scale meta-analysis study highlighted strong regional variation that makes the design of a widely active vaccine a challenge(5). A more attractive target, the O-antigen, has greatly reduced diversity(6), which might, however, be a reflection of limited surface exposure and, thus, less pressure to be kept at high diversity within the population(7). Several studies have shown limited antigenic potential of O-antigen, which is likely masked by the capsule. Assessing the best strategy for these already complex polysaccharides is further complicated because *Klebsiella* spp. are environmental bacteria, and evolutionary selection likely happens outside the human host where different pressures (in particular phage predation) impact selection(8, 9).

Proteins are, therefore, an attractive vaccine target in the case of *Kpn,* as they are often highly restricted in their ability to change due to specific functions they need to perform, such as binding to host cells, and as structures that need to be supported by the physicochemical properties of their constituent amino acids, limiting the exchangeability of residues. It has been shown in other encapsulated bacteria that not only adhesins, which reach beyond the bacterial surface for binding, e.g., to host cells, but also porins, can be recognised by antibodies, as the polysaccharide layers form channels around these, likely to enable better nutrient import(10).

An additional challenge of protein targets is that we currently have a poor understanding of which *Kpn* proteins are surface-exposed, expressed during human colonisation or infection, and, importantly, whether they can be recognised by the immune system despite the large polysaccharide capsule. Identification of targets that are expressed and recognised is a crucial step in moving towards a protein-based vaccine, as conservation across the population genome diversity must be linked with accessibility to the immune system. It is furthermore well-known that antigen profiles differ considerably geographically and between different populations, and an analysis of these must be based on the target population(11).

Here, we present a pilot study to identify maternally transferred antibodies that recognise *Kpn* proteins based on the immune profiles at time of birth in a mother-baby cohort in Malawi, a low-income African country with high neonatal sepsis rates, high levels of poverty and a low-resource healthcare setting. Here, *Kpn* sepsis after birth is a major cause of neonatal mortality. We selected a set of conserved *Kpn* surface proteins and used them on a protein microarray, assessing IgA and IgG reaction against these proteins in participants’ breastmilk, cord blood and maternal serum.

## Results

### Study design

Our study was embedded in a larger project assessing the burden of *Kpn*-associated neonatal sepsis at Chatinkha Nursery, the neonatal ward at Queen Elizabeth Hospital in Blantyre, Malawi. To understand the relationship between antibodies transferred from the mother to the baby via breastmilk or blood, we recruited a large cohort of mothers in labour to achieve a cohort of mother-baby dyads with an 80:20 ratio of babies that remained healthy and ones that developed neonatal sepsis diagnosed with *Kpn* (**Table 1**). Noting the risk of recruiting inadequate numbers of babies that developed culture-confirmed *Kpn* neonatal sepsis via this approach, we additionally recruited babies at the point *Kpn* neonatal sepsis was confirmed by neonatal blood culture. For the ‘case’ babies, six infections resulted in mortality, whereas there were three deaths in the control group, two of whom were infants that were infection-free and one of whom was an infant infected by *Pantoea* spp. Babies and mothers were screened for *Kpn* gut mucosal carriage by stool culture or rectal swab and all *Kpn* isolated, whether from blood or stool, were whole genome sequenced. Breast milk, maternal blood and cord blood (as a proxy of neonate blood) were sampled, and serum was extracted from blood. All samples were prepared for ambient temperature shipping to ADI on desiccated filter papers, where the samples were then reconstituted and exposed to the *Kpn* surface protein microarrays. A total of 69 breastmilk, 113 sera and 55 cord blood samples were probed and subsequently analysed (**Table 1**).

**Table 1.** Characteristics of the study population.

| Characteristic | Case N = 20<sup>A</sup> | Control N = 95<sup>A</sup> | p-value<sup>B</sup> |
| --- | --- | --- | --- |
| **Gender** |  |  | 0.2 |
| Female | 10 (63%) | 40 (44%) |  |
| Male | 6 (38%) | 50 (56%) |  |
| Unknown | 4 | 5 |  |
| **Age in days** | 8 (0 - 93) | 1 (0 - 77) | <0.001 |
| Unknown | 0 | 2 |  |
| **Gestational age (weeks)** | 34 (26 - 39) | 38 (24 - 41) | 0.075 |
| Unknown | 3 | 7 |  |
| **Birthweight (g)** | 2,100 (1,000 - 3,400) | 2,923 (900 - 3,800) | 0.004 |
| Unknown | 2 | 7 |  |
| **HIV status** |  |  | >0.9 |
| Negative | 16 (84%) | 76 (84%) |  |
| Positive | 3 (16%) | 14 (16%) |  |
| Unknown | 1 | 5 |  |
| **VDRL status** |  |  | 0.5 |
| Negative | 15 (94%) | 72 (96%) |  |
| Positive | 1 (6.3%) | 3 (4.0%) |  |
| Unknown | 4 | 20 |  |
| **Maternal Kpn colonisation** | 3 (15%) | 34 (36%) | 0.070 |

<sup>A</sup>n (%); Median (Min - Max). <sup>B</sup>Pearson's Chi-squared test; Wilcoxon rank sum test; Fisher's exact test.

### Genomic epidemiology of *Kpn* isolates from cases and infection outcomes

Analysis of whole genome sequence data from the isolates of the 20 neonates that developed *Kpn* sepsis revealed that they belonged to 12 different sequence types (ST). Six of the cases resulted in mortality, including 3/5 (60%) ST39 cases, 1/2 (50%) ST258 and the single ST45 and ST607 cases (**Figure 1A**). ST39 was present in our cohort with two capsule types, KL23 (two isolates) and KL62 (three isolates); all mortality-associated cases carried the KL62 operon, although another case with KL62 (ST874) survived (**Figure 1B**). The twenty isolates were predicted to have five different O-antigen types, 13 predicted as O1ab, two isolates each predicted as O3b, O2a, O2afg, and one predicted as an O-antigen loci with yet unknown serotype OL103 (**Figure 1C**). The ST39 isolates alone in this study highlight the high diversity, encoding for O1ab + KL23 (n=1), O1ab + KL62 (n=3), and O2afg + KL23 (n=1). Only one isolate carried predicted virulence factors typical for *Kpn* besides yersiniabactin; CNSJXZ, an ST45 isolate, was predicted to encode for aerobactin but none of the other virulence factors, importantly not for *rmpA2*, which causes the hyper-mucoid phenotype, were detected.

**Figure 1.**
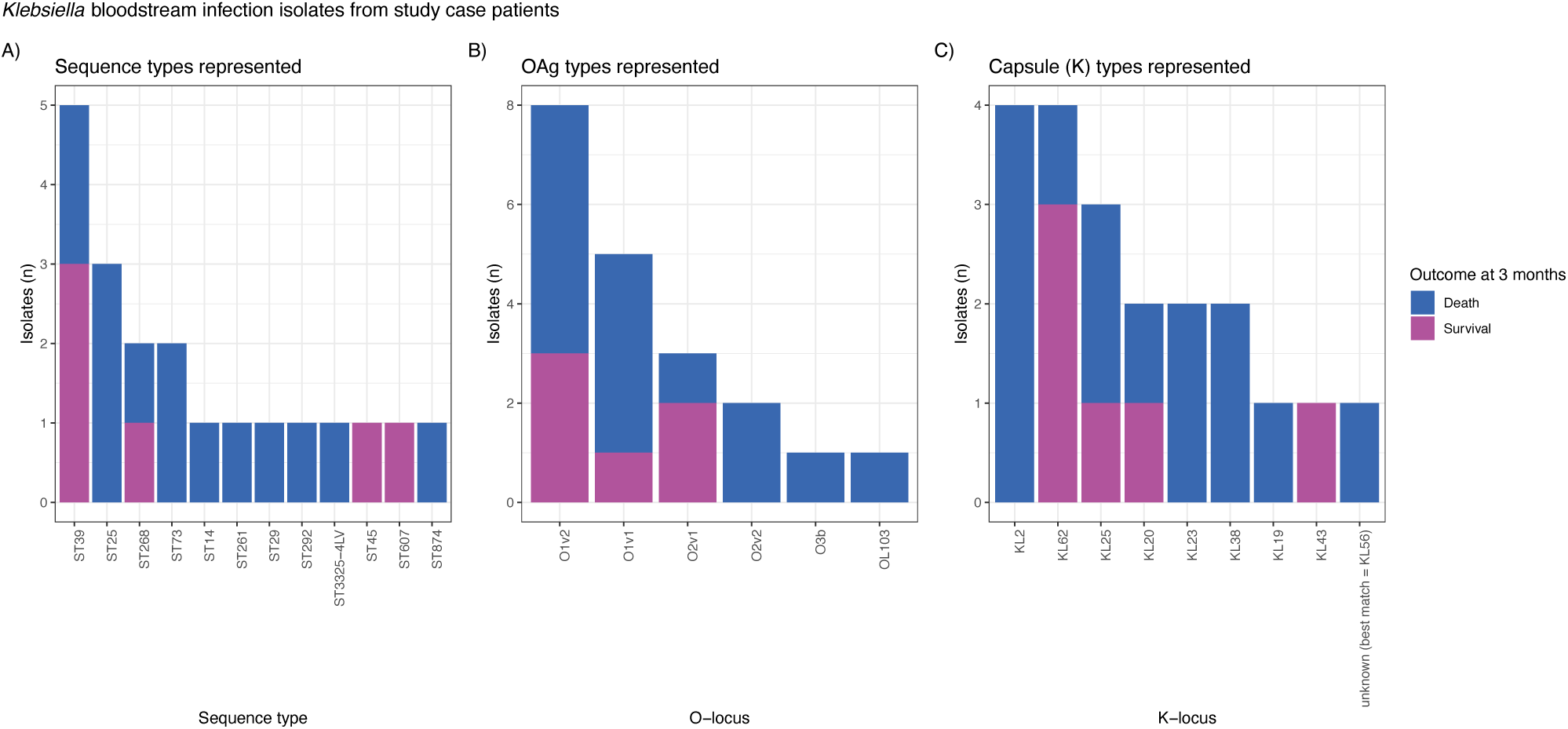
Characteristics of the *Klebsiella pneumoniae* isolates causing infection in the study cohort. (**A**) Sequence types, (**B**) LPS O-antigen types, (**C**) capsule types. Colours indicate the 3-months outcome.

### Identification of antibodies in mothers that are transferred to their babies

Approximately 10-20% of microarray *Kpn* proteins were recognized by antibodies present in at least 10% of mothers in maternal serum, cord blood and/or breastmilk (**Figure S1A-D**). A total of 33 array targets were recognised by at least one sample above the seropositivity threshold, and 30 targets were recognized by at least 10% of mothers in maternal serum IgA or IgG (10 and 19, respectively), cord blood IgG (19), and breastmilk IgA (12) (**Figure 2A**). There was substantial heterogeneity observed between mothers in both magnitude and breadth of responses (number of antigens recognized) (**Figure 2B**). Cord blood and maternal serum IgG responses were closely correlated, whereas breastmilk and serum IgA responses were less correlated (**Figure 2A**), indicating that placental transfer of IgG was strongly represented in the serum of mothers, whereas transfer of IgA antibodies by breastfeeding likely represents a more localized mucosal immune response. The proteins analysed here include several lipoproteins and components of large extracellular structures, in particular the F-type pilus and fimbriae, but also large porin-like structures (**Figure 2B**).

**Figure 2.**
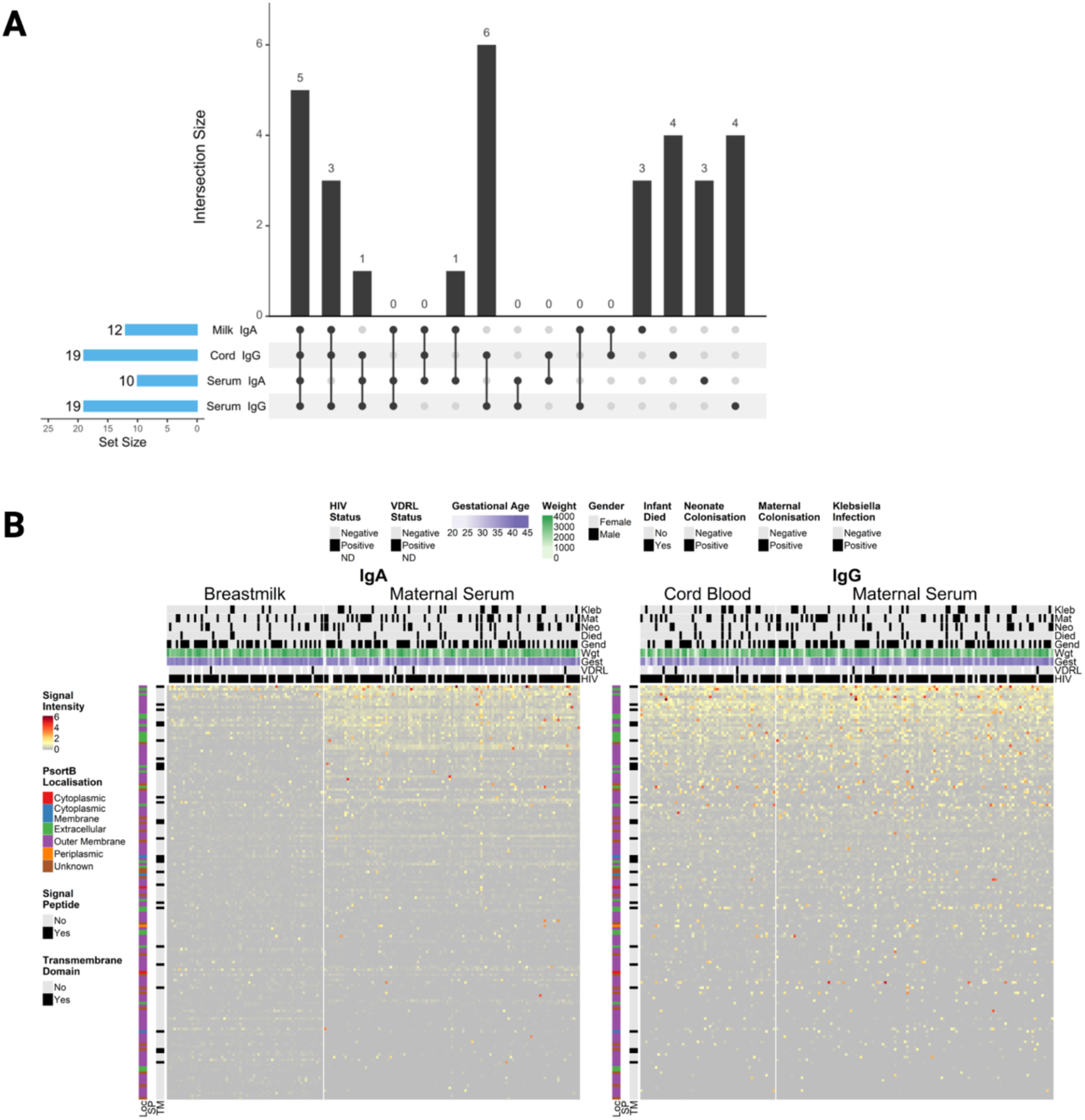
Reactivity of *Kpn* proteins with IgA and IgG antibodies in breastmilk, maternal sera and cord blood. (**A**) The upset plot shows the number of overlapping antigens between each specimen type/isotype category in the black vertical bars with the corresponding intersection between categories shown below in the dot matrix. The horizontal blue bars show the total number of reactive antigens for each category with approximately twice as many IgG responses detected. (**B**) The heatmap shows the microarray signal intensity of IgA and IgG binding on a colour scale for normalised signal intensity. Overlaid clinical characteristics are shown in column headers, and protein features are shown in row headers. Kleb: *Kpn* infection; Mat: maternal *Kpn* colonisation; Neo: neonate *Kpn* colonisation; Died: infant with *Kpn* sepsis died; Gend: gender; Wgt: weight at birth; Gest: gestational age (weeks); VDRL: syphilis test; HIV: HIV status; Loc: subcellular localisation predicted by PsortB; SP: signal peptide predicted by SignalP; TM: transmembrane domain(s) detected by TMHMM.

### Antibody responses may be associated with protection from neonatal *Klebsiella* infection and death

To explore factors associated with antibody binding to *Kpn* antigens, multivariable linear models were fit. There was no apparent relationship between available clinical and demographic variables on most antibody responses to *Kpn* antigens recognised in maternal breastmilk, serum and cord blood (**Figure S2**). These variables included maternal HIV status, syphilis/VDRL test status, *Kpn* colonisation, birthweight, gestational age, gender and age in days at recruitment.

Univariate analyses were performed to compare antibody responses between infants with culture confirmed *Kpn* neonatal sepsis and controls. After correction for the false discovery rate, only a few antibody responses remained significantly different, although the differences in means were small. However, the upper quartile of responses in the uninfected group were notably higher than responses in the upper quartile of the *Kpn* infected group and may represent a subset of strong antibody responders that are able to resist bloodstream infection (**Figure 3**). Importantly, we cannot distinguish individuals in the uninfected group that remain uninfected due to immunity from individuals that remain uninfected due to lack of exposure. Multivariable analysis using logistic regression agreed with univariate analysis, with most *Kpn* protein antibody responses trending to protective effects but non-significant after correction **Figure S3**.

**Figure 3.**
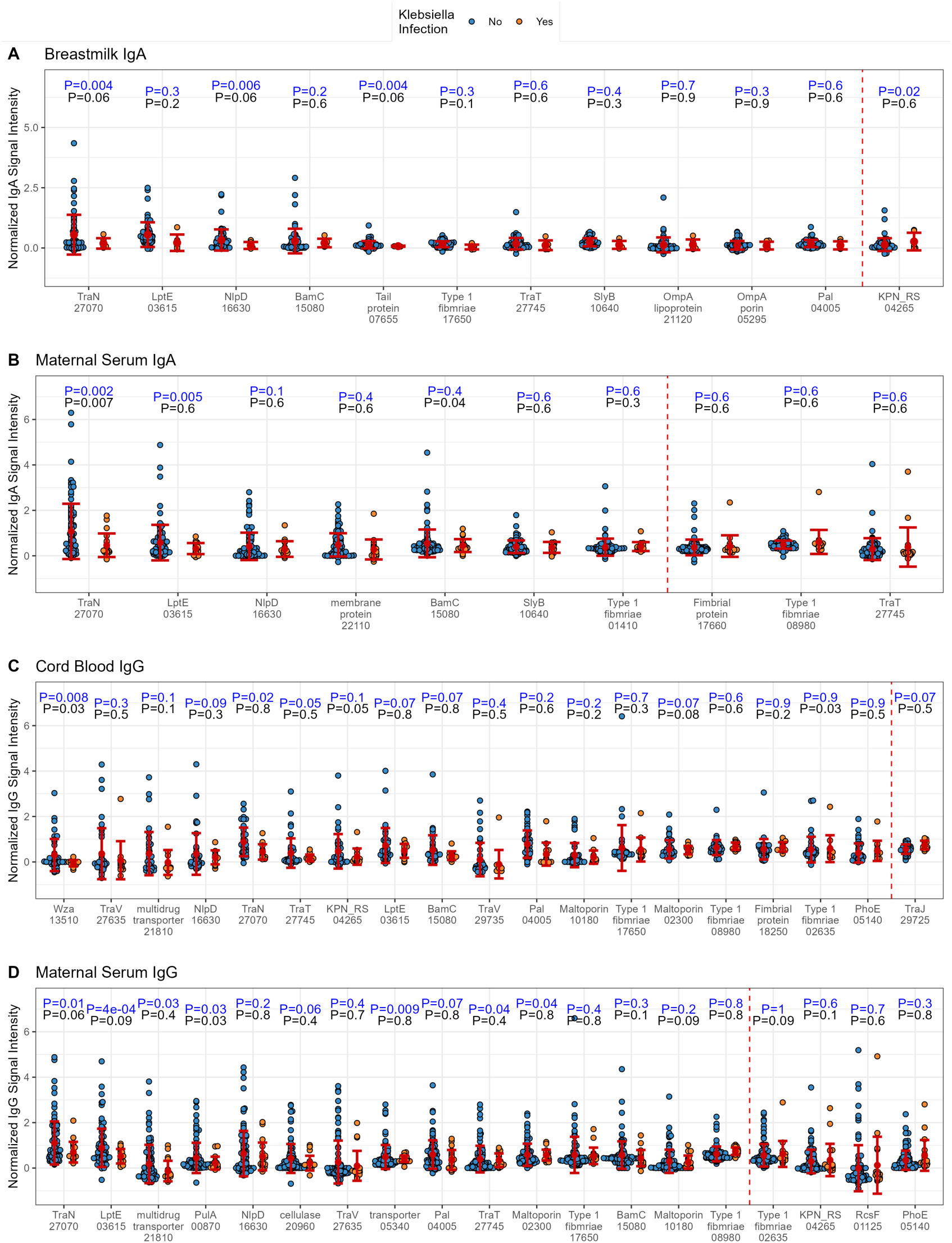
Differential antibody reactivity to Kpn proteins between infant cases of Kpn sepsis and non-case controls in breastmilk, maternal sera and cord blood. The beeswarm plots show the microarray signal intensity distribution of individual data points for each reactive antigen for (**A**) breastmilk IgA, (**B**) maternal serum IgA, (**C**) cord blood IgG and (**D**) maternal serum IgG. Blue points represent infant non-case controls, and orange points represent infant cases of *Kpn* sepsis. Red points represent group means with error bars representing the standard deviation. Individual proteins are shown on the x-axis with protein name and RefSeq gene ID tag (5 digits following “KPN_RS”). T test P-values corrected for the false discovery rate for each comparison are shown in black above each protein. Notably, subjects in the upper quartile in the uninfected group tended to have higher signals than the infected group, thus additional T tests were performed on upper quartiles of each group and P-values are shown in blue. Antigens are ordered by mean differences in the upper quartile of each group. The vertical dashed line partitions the antigens with higher upper quartiles in the uninfected group (left of the line) and those with higher upper quartiles in the infected group (right of the line).

In human milk, notable proteins with higher upper quartile responders in the uninfected group were the conjugation machinery protein, TraN, the Tail protein (phage tail length tape measure family protein), the murein hydrolase activator, NlpD, LPS assembly lipoprotein LptE, and the outer membrane protein assembly factor, BamC. Whilst maternal serum IgA responses to some proteins such as TraN, BamC, LptE, NlpD and other membrane and fimbrial proteins had more responders in the uninfected group, it is unclear how maternal IgA signals may relate to protection in infants. In the cord blood and maternal serum, IgG to LptE, BamC, type 1 fimbriae, Pal and NlpD, type IV conjugative transfer system proteins TraN, TraV and TraT, the pullanase protein PulA, multidrug transporter, and cellulase and maltoporin proteins had higher upper quartile responses in the uninfected group. In cord blood IgG, responses to polysaccharide export protein Wza were also serorecognized in more samples linked to uninfected infants. Among *Kpn*-infected infants that died (n=6), no antibody responses were associated with death (**Figure S4**).

To assess if multiantigen profiles were associated with having neonatal *Kpn* infections or not, all IgA and IgG responses to reactive antigens in each body fluid type were included in a partial least squares discriminant analysis (PLS-DA). While there was substantial overlap in a subpopulation of mothers paired to both infected infants and infants with no *Kpn* infections, the two groups trended in opposite directions within the first component, X-variate 1 (**Figure 4A**). Analysis of the model variables showed that numerous antigens were enriched in the population of mothers with children that remained uninfected during follow-up, even though some antigens were associated with higher odds of getting infected. The proteins explaining the majority of the signal are part of the conjugation machinery (TraJ, TraT, TraN), large extracellular structures including fimbriae, proteins that are likely to be highly expressed like the BAM machinery and the LPS secretion system (BamA/K, LptE) but also lipoproteins (LysM peptidase, OmpA) and classical porins and efflux proteins. These data suggest that a subset of antigens may be implicated in protection, while another subset of immunoreactive antigens may be associated with increased risk of infection, or perhaps increased exposure. The most important responses in the model were serum and cord blood responses, although both IgA and IgG were represented (**Figure 4B**). Although underpowered for multivariable analysis with clinical characteristics in this heterogeneous population, an analysis of the top antigens in linear models demonstrated a trend for the strongest responders to be in the uninfected group (**Figure 4B**).

**Figure 4.**
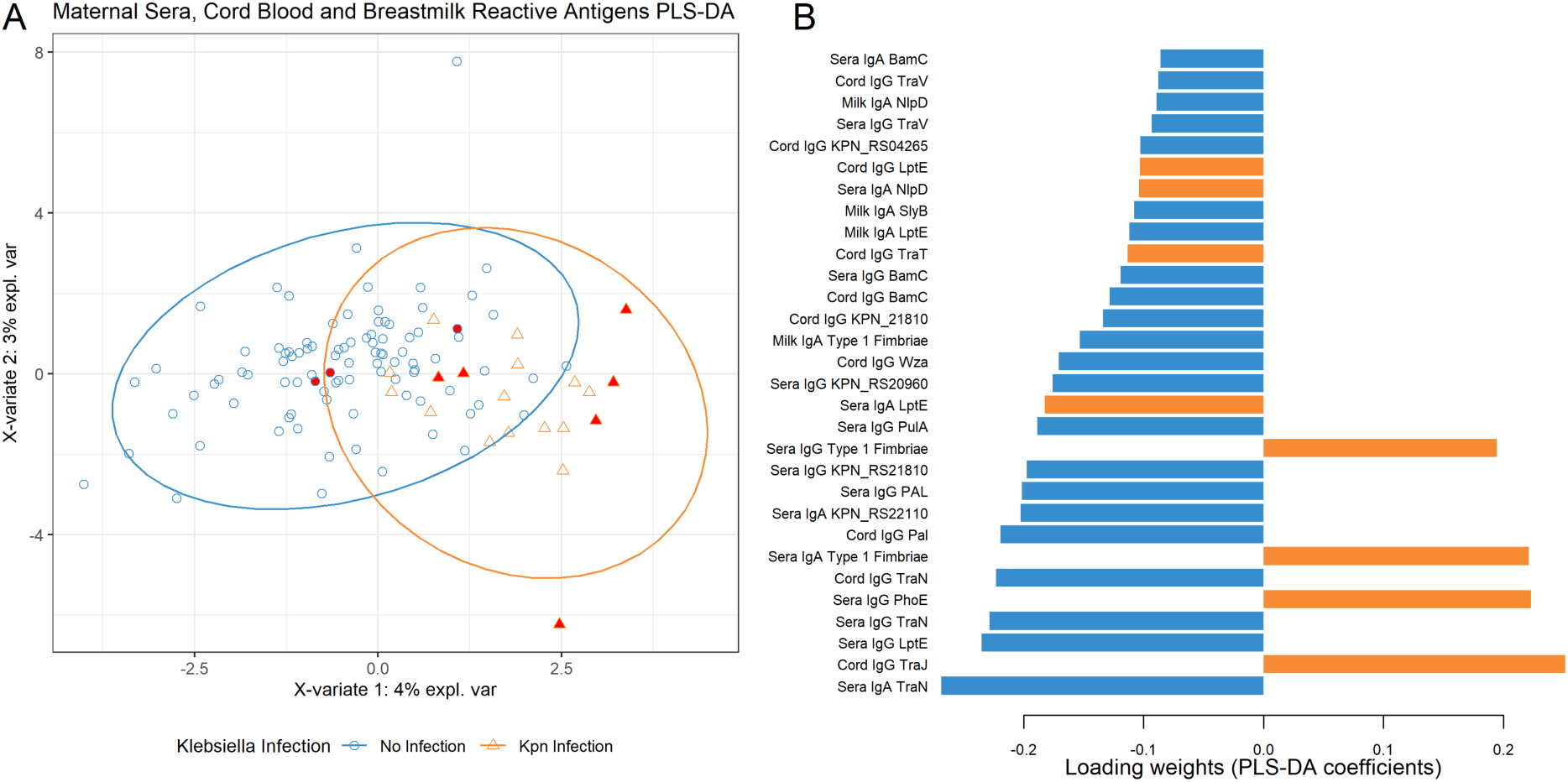
Partial Least Squares Discriminant Analysis (PLS-DA) shows unique antibody reactivity profiles in the population of mothers with infants that had *Kpn* infections vs. those that did not. (A) The score plot shows the distribution of all samples and antibody isotype measurements in the PLS-DA model. Orange triangles represent samples associated with infants that had *Kpn* infection, and blue points represent samples associated with infants without *Kpn* infection. Red-filled triangles represent infant *Kpn* sepsis cases that died. Ellipses are drawn around the distribution of each group. (B) The loadings bar plot represents the contribution of each antigen measurement to the principal components of the PLS-DA model. Bars at the bottom, furthest from 0.0 represent the greatest role in separating *Kpn* infected from non-infected infants. Blue bars represent responses enriched in the non-infected group, and orange bars represent responses enriched in the *Kpn*-infected group.

### Large surface structures and lipoproteins from large surface complexes dominate high signals

The target selection for the chip design focused on known outer membrane-associated protein profiles rather than specific genes, thus capturing an insight into reactions of paralogues (homologous copies of the same protein) and larger protein families (like multiple distinct fimbrial operons), enabling us to compare reactivity of different structures as well as closely related targets. The highest reactivity overall was observed in proteins forming large extracellular structures, including outer membrane components of the conjugation machinery (TraN, TraV, TraT) essential for F-plasmid conjugation(12, 13), as well as several fimbrial subunit proteins, which form part of large fimbrial structures by forming a stalk-like extension of interlinked domains(14). However, only 8/29 fimbrial subunits on the microarray were recognised in this cohort, despite their structural similarity and equal involvement in large surface structures, and none of the 11 usher proteins (**Figure 5A**), raising the importance of assessing the full fimbrial diversity, conservation and expression patterns, across the complex pangenome of *Kpn*. Despite concerns about the large capsule, several large porins and other beta-barrel structures were identified as reactive (OmpA, LamB-like, PhoE, Wza). We further noted that small lipoprotein components were detected (15/30 targets with significant signals have a predicted lipid anchor). Several of these represent lipoprotein components of larger complexes, including LptE, part of the LptDE lipopolysaccharide export complex(15), and BamC(16), part of the BAM complex. In addition, several identified lipoproteins are known to be found adjacent to outer membrane proteins (SlyB(17)) or involved in processes that might lead to greater exposure (e.g., Pal and NptE(18), involved in cell division at the site of septum formation). Considering the presence of the highly reactive proteins in *Kpn* circulating in our cohort, pangenome analysis of the 20 isolates from our study cases confirms that the majority (19/30) were conserved in all isolates (**Figure 5B**), 5/30 were conserved in >10 of the genomes, and 6/30 present in <10 of the genomes in our pilot study cohort.

**Figure 5.**
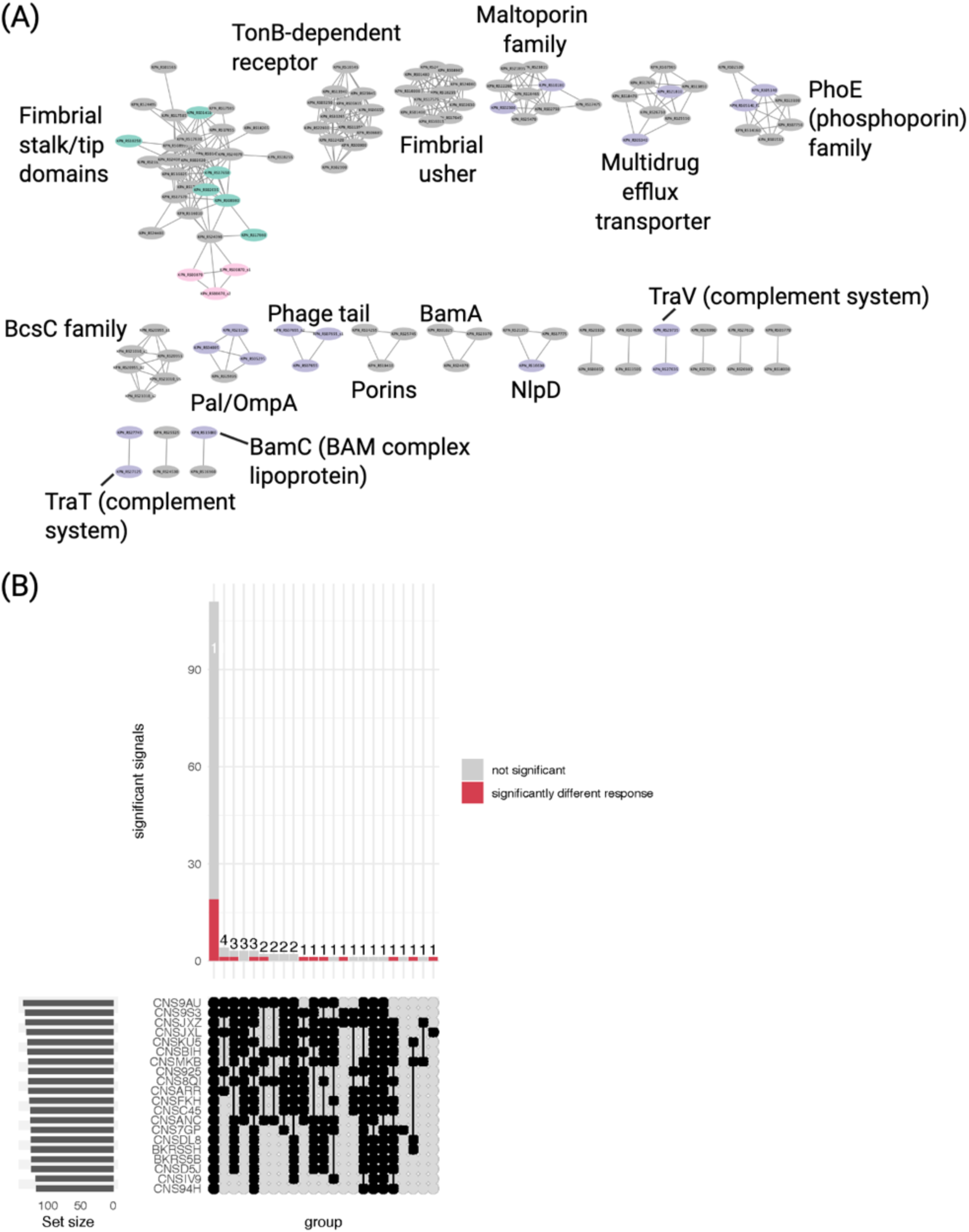
Similarity network of all sequences represented on the protein microarray and conservation within study population clinical *Kpn* isolates. (A) The protein-protein sequence similarity network shows coloured proteins with antibody reactivity. Violet: outer membrane; green: extracellular; pink: unknown. The green and pink data edges are part of the large fimbrial domain cluster. (B) The UpsetR plot of all genes on the chip and their conservation in the 20 *Kpn* isolates from case infants. The red colour indicates proteins that were reactive to IgG or IgA antibodies in breastmilk, maternal sera or cord blood. A total of 18 were perfectly conserved, 5 were conserved in most (>50%) and 6 in <50%, i.e., poorly conserved.

## Discussion

We investigated antigen reactivity against a protein microarray of 161 different *Kpn* proteins or protein fragments representing 152 unique proteins predicted to be surface-exposed or secreted in a cohort of mother-baby dyads in Blantyre, Malawi. This study represents a first, essential step towards developing a *Kpn*-targeted protein vaccine or monoclonal antibody treatment. Given the large capsular polysaccharides surrounding the cells, it could be speculated that similar challenges for protein antigens exist as for LPS O-antigens, where the capsule has been noticed to shield antibody-antigen interactions(7). However, in this case, we see antibody reactivity even against proteins likely to be cell surface associated (such as porins and outer membrane lipoproteins), which might be due to the need for their accessibility for nutrient import and export(10). We furthermore observed high reactivity against large extracellular structures, in particular the conjugation machinery and fimbriae. These represent attractive vaccine targets, as they are structurally restricted, i.e., vaccine escape mutants would likely lose the protein functionality, and represent key factors of *Kpn* that drive its problematic nature as an ESKAPE pathogen, in particular the acquisition of resistance plasmids from other cells via the conjugation machinery.

The antibody profiles of the samples linked to uninfected infants suggests that there may be protective antibody responses, but more data is needed to support this finding. In particular, the uninfected infants represent a heterogeneous population, including both those that may not have been significantly exposed to *Kpn* but remain at risk from *Kpn* neonatal sepsis, and infants that might have passively received maternally derived protective antibodies resulting in asymptomatic exposure to *Kpn*. Additionally, the lack of associations with potential confounding factors and loss of significance after correction for the false discovery rate, despite plausible targets of immunity, indicate that this study lacked power for these multivariable analyses at the dimensionality of protein microarrays. Nevertheless, valuable leads were identified in the data, including identification of both passively transferred systemic (serum/cord IgG) and mucosal (milk IgA) antibody targets. Maternal *Kpn* colonisation seems to play a minor role in passive transfer of antibodies, likely eclipsed by antibodies accumulated through lifetime exposure, but also indicating that colonisation in itself may not cause elevated antibodies in an exposed adult population.

Milk IgA antibodies showed a similar trend to cord and maternal serum IgG for some antigens, indicating the potential role of breastfeeding in protection from *Kpn* infection, as has been observed for other pathogens(19–27). Although maternal serum IgA is unable to be passively transferred to the infant, these responses may reflect the antibody repertoire present in breastmilk, also supported by the similar responses to TraN, LptE and BamC between maternal serum and milk IgA. This raises the possibility that some antigens could be targets for oral vaccines or other mucosal immunity-directed vaccines, particularly if maternal vaccination could translate to high levels of specific milk IgA for passive transfer during breastfeeding. Alternatively, these responses may simply be colinearly associated with the maternally transferred IgG response, as the same antigens were associated with lack of infection in maternal serum and cord blood IgG.

It seems that large extracellular structures like fimbriae and the conjugation / plasmid transfer machinery, and abundant complexes like BAM (BamC) and LptDE (LptE), are strongly recognised, which might be a combination of their prevalence and exposure. Whilst fimbriae and pili clearly extend beyond the capsule, the large membrane-embedded complex structures likely form a channel in the LPS and polysaccharide as observed for *Salmonella*(10). It is notable that the small lipoprotein components of the larger complexes, BamC and LptE, were particularly reactive. Whilst we cannot exclude an impact of better folding of these and thus greater likelihood of presentation on the microarray in native conformation, we observed signals for several large beta-barrel structures (OmpA, maltoporins), indicating that large membrane-embedded proteins can be recognised. These responses also suggest that lipoproteins might be more surface exposed at times. BamC surface exposure has previously been highlighted(16), and the location of LptE in the LPS export machinery within the barrel of LptD already suggests easy surface recognition(28).

Following WHO recommendations, the antenatal care (ANC) plan in Malawi has been updated to 8 visits(29). There are various efforts to improve adherence to these, which is of high interest for positive pregnancy outcomes and reducing maternal mortality overall(30). For specific cases like unvaccinated mothers, the tetanus toxoid-containing vaccine is recommended during pregnancy in LMIC settings, further emphasizing that an introduction into the antenatal care plan may not need implementation of novel routines but could be linked to already ongoing efforts, although optimal timing of maternal *Kpn* vaccine administration would need to be determined, given that prematurity is a major risk for neonatal sepsis.(31, 32).

This study identified conserved, highly immuno-reactive proteins, as well as observing initial indications for protective antibodies, thus forming a foundational evidence base for further investigations into a protein-based maternally-administered vaccine against neonatal sepsis caused by *Kpn.* The data presented provide new leads for hypotheses about specific antibodies that may prevent *Kpn* sepsis. Though not all infants in the uninfected group had elevated *Kpn* antibodies, and it is unknown which had been exposed to *Kpn*, the antibodies seen in a strongly reactive subset of the mothers with infants that remained uninfected should be further evaluated as correlates of protection that may serve as candidate antigens for vaccines and therapeutics to protect infants from the significant morbidity and mortality of *Klebsiella* infections.

## Methods

### Sex as biological variable

As the study was based on mother-infant dyads, with samples collected from the mother and cord blood at time of delivery, samples were derived only from female participants, and sample selection was not based on the sex of dyad infants. However, sex was included as a variable in all multivariable models to assess the effect of infant sex on antibody responses, as well as adjustment of other covariate effects.

### Clinical setting

Queen Elizabeth Central Hospital (QECH) provides healthcare free at the point of delivery and is the tertiary healthcare centre for the city of Blantyre and referral centre for all southern Malawi. Chatinkha nursery is the neonatal unit at QECH; there are approximately 14,000 neonates born and 5,000 admissions to the Chatinkha nursery at QECH each year. Approximately 80% of babies are born inside QECH, and *Kpn* neonatal sepsis is a major healthcare concern(33); overall neonatal mortality in the unit was 17% in 2013(3).

### Study design

This study was a case-control nested within a prospective cohort study. Mother-infant pairs were eligible to be recruited into the study if they met one of the following two criteria: i) mothers in labour who were expected to deliver at QECH; ii) mother-infant pairs of infants less than three months old on the paediatric wards at QECH. We specifically recruited mother-infant pairs from the paediatric wards where the infant had infection with *Kpn* identified by its presence in a normally sterile site (blood or CSF), however, mother-infant pairs where there was no infection in the infant were also recruited. These cases were identified as per the microbiology laboratory below. In the case of twins, both were recruited. This resulted in total recruitment of 908 mother-infant pairs in total from April 2021 – April 2023, of which 637 were recruited in labour and 271 were recruited from the paediatrics wards. 154 of these developed clinical signs of sepsis, and 24 of these had *Kpn* infection. From these, we selected 80 controls with no *Kpn* infection and 20 cases with *Kpn* infection. Outcomes were recorded up to three-months postnatal age.

Dyads recruited in labour had serum sampled prior to delivery and cord blood sampled immediately post-partum as a proxy for the infants’ serum. For dyads recruited on the paediatrics wards, maternal serum and infants’ serum was sampled (no cord blood was available). Maternal breast milk (colostrum) was also sampled post-partum when available and frozen at -80°C immediately after sampling. A protocol was developed for collection of samples on Whatman filter paper, and peripheral blood spots, cord blood spots and breastmilk spots were collected and shipped for subsequent protein microarray analysis. Milk samples were thawed at room temperature and homogenised by pipette mixing. To prevent mold growth on filter papers with milk samples, a 5% sodium azide solution was added to milk to a final concentration of 0.02% prior to adding milk to filter papers. For all samples, a volume of 50µL was spotted onto 1cm x 2cm cut filter paper folded into a 2mL screwcap tube, dispensing slowly to the top of the filter paper and allowing the volume to permeate to the bottom. Filter papers were left to dry overnight at room temperature, and tubes were closed once filter papers were dry and kept at room temperature until shipment to ADI.

### Surface protein selection

To identify the most likely surface proteins for *Kpn*, a comparative analysis of a core dataset was performed(34). These proteins were then searched for a list of surface domains predicted based on Pfam profiles, which were selected initially as being members of the CL0193 MBB clan as initial core selection. These profiles were then assessed using both HHSearch(35) results and manual assessment of reciprocal below-cutoff hits, and Pfam profiles that were similar to profiles in the MBB CL0193 clan were assessed manually and added to CL0193 if appropriate. Domains associated with these putative outer membrane proteins (for example adhesin domains) were collected as well and assessed for inclusion. This resulted in a library of 225 Pfam domains likely surface-exposed, outer membrane embedded, or both, that were used as search input against the *Kpn* genome collection (Table S1). The domains were defined as “membrane anchor” if resembling a beta-barrel-like structure and predicted to be embedded in the outer membrane, “extracellular profile” if secreted or parts of extracellular domains (for example Ig-like adhesin domains), and “other secretion” to label other secretion systems that differ structurally from classical OM adhesins or porins, such as the type 2 secretion system. Additional domains identified in the selected proteins were broadly grouped into “repeat”, for example tetratricopeptide repeat domains, “small” to group domains often associated with large membrane proteins such as the plug domain of TonB-like receptors or N- or C-terminal conserved domain of large membrane structures, “binding / enzyme” to denote domains that are predicted to bind co-factors or function enzymatically (e.g., protease domains), “lipo” for domains describing lipoprotein families, and “inner membrane” for domains resembling transmembrane helices; see **Table S1** and **Figure S6**.

### Protein domain analyses

The proteins were mapped back to Pfam domains(36) using InterProScan (v104)(37), signal peptides predicted using SignalP (v6.0)(38), and visualised using iToL(39). A pairwise similarity based network was constructed using the CLuster ANalysis of Sequences (CLANS) software(40), which uses a version of the Fruchterman-Reingold graph layout algorithm to visualise pairwise sequence similarities. The program was used at default settings for 2-dimensional clustering and the clustering repeated >10 times to ensure capture of a representative image of the cluster layout. The sequence similarity network visualisation was exported directly from CLANS, and the network information was exported for import into cytoscape (v3.10.3)(41) for additional analyses overlaying the detection signals measured as above.

The protein microarray contained 161 unique protein fragments; one of these included one additional and four of these two additional shorter variants to account for potential accessibility issues given their extensive length, representing 152 unique *Kpn* proteins (**Figure S5**). The protein microarray represented 82 distinct domain architectures of 95 distinct domains and 4 proteins with no Pfam domains identified but other indicators for potential surface exposure (signal peptide prediction, membrane location motifs) (**Figure S6)**. The Pfam domains detected in the proteins include 82 targets with predicted membrane anchor profiles (likely embedded in the outer membrane, with potentially surface-exposed loops or other domains), 29 with domains likely exposed to the extracellular space (e.g., domains frequently found in adhesins), 12 other secretion system profiles, where each protein can encode for multiple domains of similar or combinations of categories as above (**Figure S6, Table S2)**. 142 proteins were likely secreted based on a signal (106) or lipo-signal (36) peptide. Of the 82 surface-anchored targets, 45 are likely membrane-embedded with only loops or short extensions accessible from the outside (i.e., encoding only membrane-anchor profiles), whilst 37 are putative larger adhesive structures with surface-exposed components (i.e., encoding a membrane anchor as well as an additional domain).

### Microarray Development

The list of prioritized *Kp* proteins was developed as above and provided to ADI. This list included predicted surface structures and was enriched for predicted outer membrane proteins, proteins with signal peptides and proteins with transmembrane domains. The protein microarray was prepared as described previously(11); in brief, genes were cloned at ADI into ADI’s expression vector and used for high throughput cell-free in vitro transcription and translation. The open reading frames were subcloned into a T7 expression vector pXI and expressed using an in vitro cell-free *Escherichia coli* transcription and cell-free translation (IVTT) system (Rapid Translation System, Biotechrabbit, Berlin, Germany). After expression, the proteins were printed onto nitrocellulose-coated AVID slides (Grace Bio-Labs Inc) using an Omni Grid Accent robotic microarray printer (Digilabs, Inc., Marlborough, MA, USA). In total, 161 *Kpn* proteins and protein fragments were printed onto microarrays.

### Probing and quantification of antibodies in maternal plasma, cord blood and breastmilk samples

Antibodies were eluted from the shipped filter papers by adding 500µL of Surmodics Assay Buffer and resuspended overnight at 4°C on an orbital shaker. Samples were pre-incubated with IVTT reaction mixture to reduce background reactivity against the expression mixture, and they were then probed on the array at final concentrations of 1:100 for maternal serum and cord blood, and at 1:15 for breastmilk. Samples were incubated on the microarrays overnight at 4°C on a rocker, subsequently washed and incubated with goat anti-human IgG-Fc fragment DyLight650 (Bethyl Laboratories, Cat#A80-104D5) or Cy3 AffiniPure F(ab’)₂ fragment goat anti-human serum IgA, α chain specific (Jackson ImmunoResearch Laboratories, Cat#109-165-011) for one hour at room temperature on a rocker, then washed, dried and stored in the dark until scanning. The exposed microarrays were scanned, and the spot and background signal intensities (SI) were exported into R for statistical analysis. Spot SI values were floored to 1. Next, the data were normalized by dividing the *Kpn* protein spot values by the median of all IVTT spots on the microarray, including IVTT control spots (IVTT expression reactions with no *Kpn* ORFs), and values were log transformed using the base-2 logarithm. Thus, normalized data represented the log_2_ signal-to-noise ratio, where a value of 0 represents specific antibody SI equal to the background, 1.0 represented twice the background, 2.0 represented 4-fold over background, etc.

### Invasive isolate microbiological processing

One to two millilitres of blood was taken from neonates (up to 28 days old) with risk factors or a clinical suspicion of sepsis. Blood was collected and inoculated and incubated using the automated BacT/Alert system. Positive samples were identified by Analytical Profile Index (bioMérieux). Antimicrobial susceptibility testing was determined by the disc diffusion method (Oxoid, United Kingdom) according to EUCAST guidelines. *Kpn* isolates were stored at -80°C.

### Sequencing of isolates from babies developing sepsis

DNA was extracted using the QIAsymphony machine and QIAsymphony DSP kit with onboard lysis, according to the manufacturer’s instructions. Quality control was done using Qubit and samples with a DNA volume of less than 200 ng were repeated. Samples that passed QC underwent Whole Genome Sequencing (WGS) at the Wellcome Sanger Institute at 364 plex on the Novaseq SP generating 150bp paired-end reads.

Isolates were recovered from the MLW sample archive and total nucleic acids were extracted using the MasterPure complete DNA and RNA Purification Kit (Bioresearch Technologies, United Kingdom) according to the manufacturer’s instructions. The normalised DNA was used for library preparation using the rapid barcoding kit (SBQ-RBK004, Oxford Nanopore Technologies plc) following the manufacturer’s instructions. The prepared library was sequenced on an Oxford Nanopore r9.4.1 flow cell on the MinION Mk1C sequencer (Oxford Nanopore Technologies plc). Data acquisition was done by the MinKNOW^TM^ software (version 20.10.6, Oxford Nanopore Technologies plc). Base calling and demultiplexing using Guppy Basecalling Software, Oxford Nanopore Technologies plc. (version 6.0.7). We used Guppy with the default settings and the “dna_r9.4.1_450bps_sup.cfg” configuration file for the guppy_barcoder. We used the command line argument “--trim_barcodes” for the debarcoding step using guppy_barcoder. For guppy_barcoder we passed “SQK-RBK004” to the “--barcode_kits” command line argument.

### Genome and pan-genome analyses

Genome assemblies were performed using a pipeline starting with flye (v2.9.2)(42). The assemblies were then polished with medaka (v1.8.0, https://github.com/nanoporetech/medaka) which uses the long read sequence data followed by polypolish (v0.5.0)(43) including the insert size filter step as recommended by the author; or unicycler(44); depending on coverage of the long reads, accessions are provided in Table S3. The assemblies were then annotated using prokka(45), and the pan-genome was calculated using panaroo (v1.3.4)(46) with the –clean_mode strict and --remove-invalid-genes flags. To include the proteins represented on the chip, we also used the *Klebsiella pneumoniae* subsp. *pneumoniae* MGH 78578 genome sequence which was used as reference genome for the genes cloned onto the protein chip; (NC_009648.1, NC_009649.1, NC_009650.1, NC_009651.1, NC_009652.1, NC_009653.1), the sequences were retrieved from GenBank and converted with the convert_refseq_to_prokka_gff.py script for compatibility with panaroo. Sequence type, capsule type(4) and LPS O-antigen type(6) were predicted using kleborate (v3.0)(47). Data was visualised using R(48), cytoscape(41) and iToL(49).

### Study approval

Ethical approval for this study was granted by the University of Malawi College of Medicine Research Ethics Committee (COMREC; study number P.10/18/2499) and Liverpool School of Tropical Medicine Research Ethics Committee (study number 19-018). The research conformed to the principle of the Helsinki Declaration.

## Data availability

Sequence data is available at SRA/ENA under bioproject PRJNA1428055; a detailed overview of accession numbers is given in Table S3. All reported data is available in the Supplementary Data file, and underlying data for all figures are included in the Supporting Data Values file. All software versions and input commands are documented in the methods.

## Author contributions

Conceptualization: JJC, NAF, EH

Data curation: JE, ADS

Formal analysis: JJC, EH

Funding acquisition: JJC, NAF, EH

Investigation: JJC, OP, AMZ, AAT, JVP, JME, ADS, LG, SL, DL, EH

Methodology: OP, AMZ, AAT, JVP, JME, ADS, SL

Project administration: JJC, EH

Resources: AMZ, PS, ET, KK, NAF

Software: JCC, EH

Supervision: JJC, OP, SL, KK, NAF, EH

Validation: OP, AMZ, PS, ET, AO

Visualization: EH, JJC

Writing – original draft: EH, JJC

Writing – review & editing: JJC, NAF, EH

All authors read and approved the final manuscript.

## Acknowledgments

We thank the Wellcome Sanger Pathogen Genomics team for expert technical support and Alexander Bateman (EBI, Cambridge, UK) for support with the Pfam domain selection.

## Supplementary figures and tables

**Figure S1.**
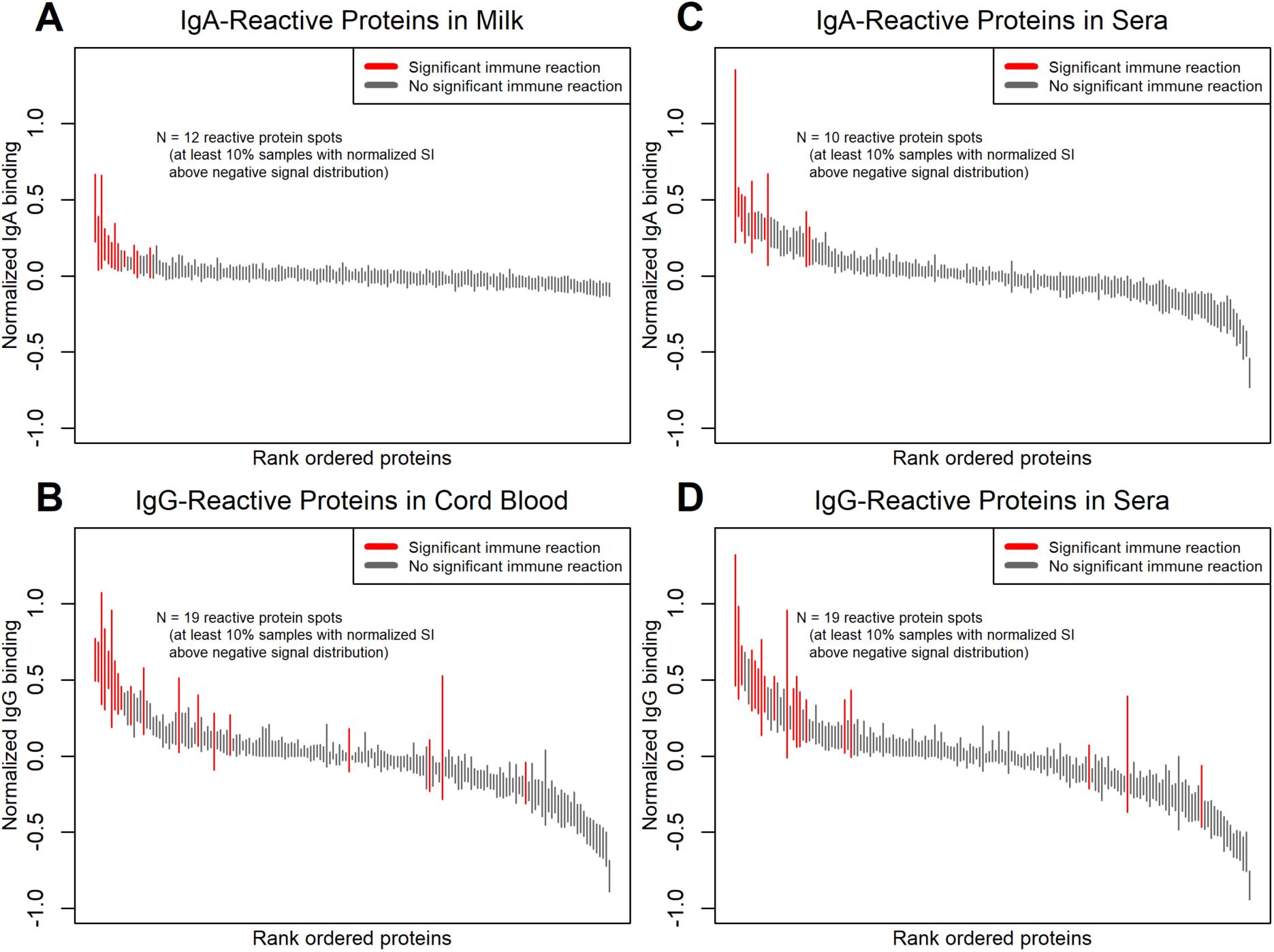
Antibody reactivity of antigens on the *Kpn* protein microarray. The errorbar plots show proteins ordered by median signals and highlighted red if at least 10% of mothers had seropositive responses for IgA in breastmilk (A) and maternal sera (C), and IgG in cord blood (B) and maternal sera (D). Bars represent the interquartile range of antibody signal intensity for eat *Kpn* protein.

**Figure S2.**
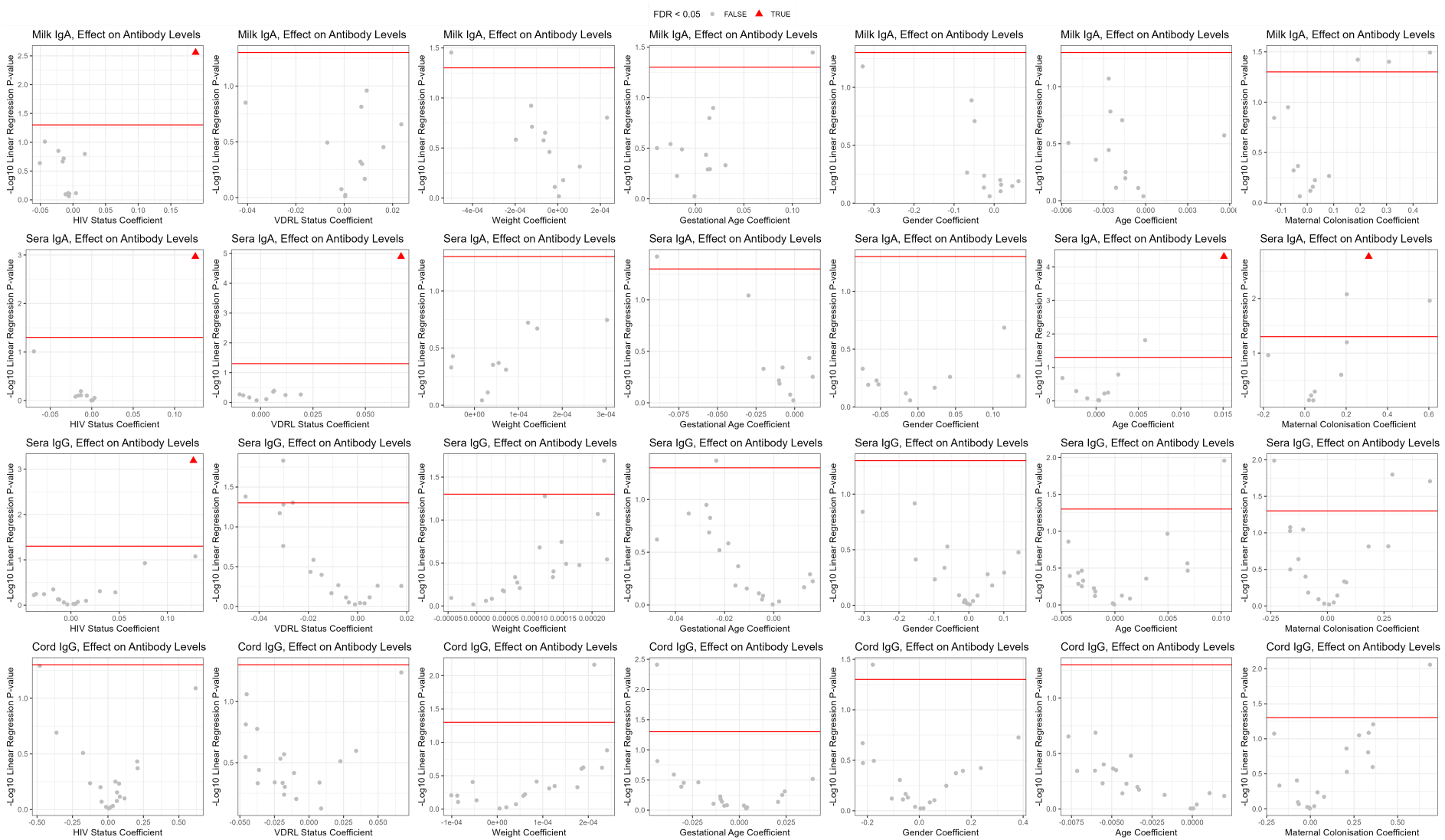
Multivariable linear models show minimal effect of clinical and demographic variables on antibody levels. The volcano plots show the multivariable ordinary least squares linear regression model coefficients on the x-axis and the inverse log10 linear regression P-value on the y-axis. The horizontal red lines represent an unadjusted P-value of 0.05, while points shown as red triangle represent differential antibody levels that were significant after correction for the false discovery rate (FDR). Each column of panels represents a model covariate, and each row of panels represents a specimen type and isotype measurement.

**Figure S3.**
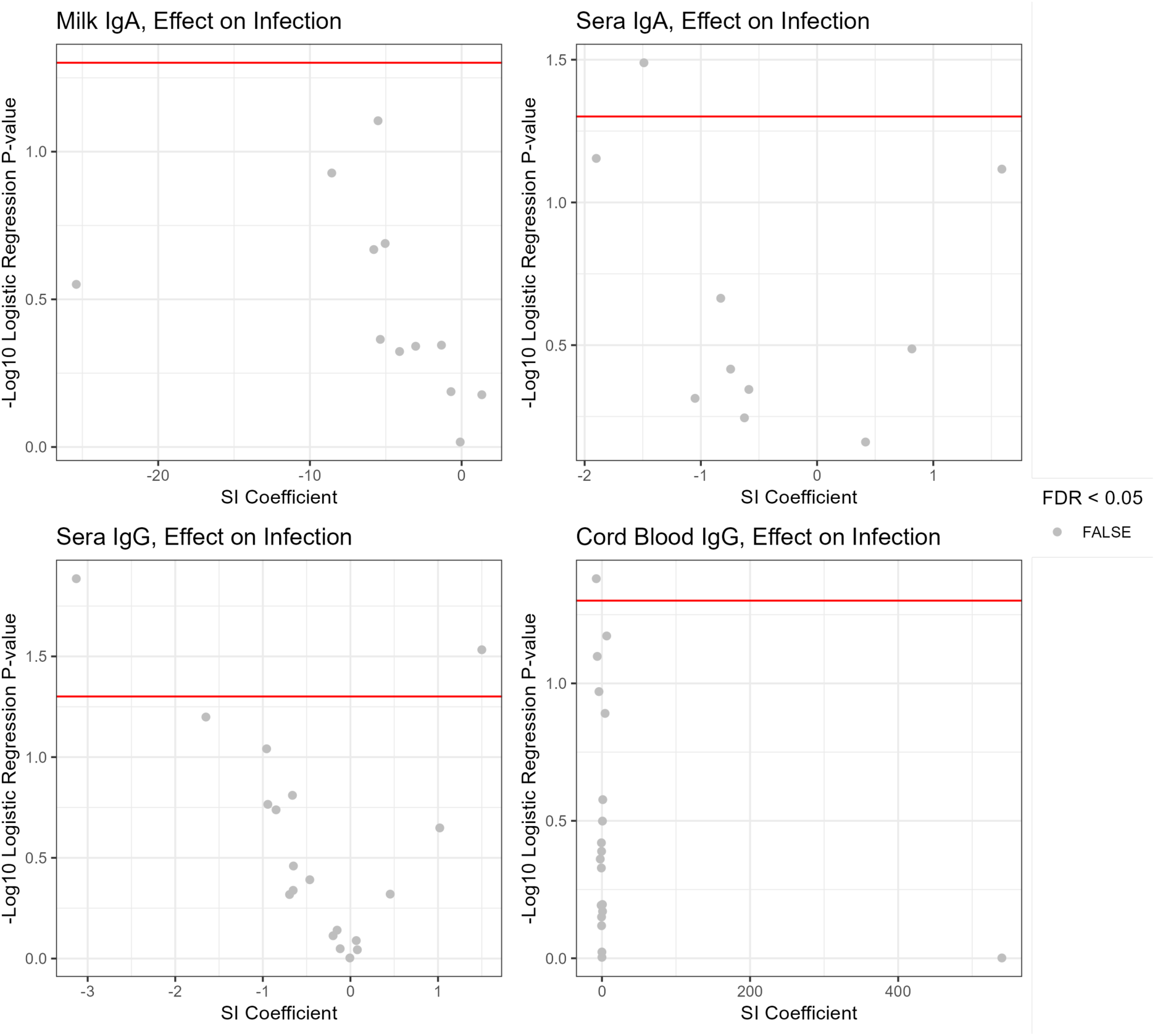
Logistic regression models show no significant effects of antibody levels on *Kpn* infections. The volcano plots show the multivariable logistic regression model coefficients of association of antibody levels with odds of infant *Kpn* infection on the x-axis and the inverse log10 logistic regression P-value on the y-axis. The horizontal red lines represent an unadjusted P-value of 0.05, while points shown as red triangle represent differential antibody levels that were significant after correction for the false discovery rate (FDR). Each panel represents a specimen type and isotype measurement.

**Figure S4.**
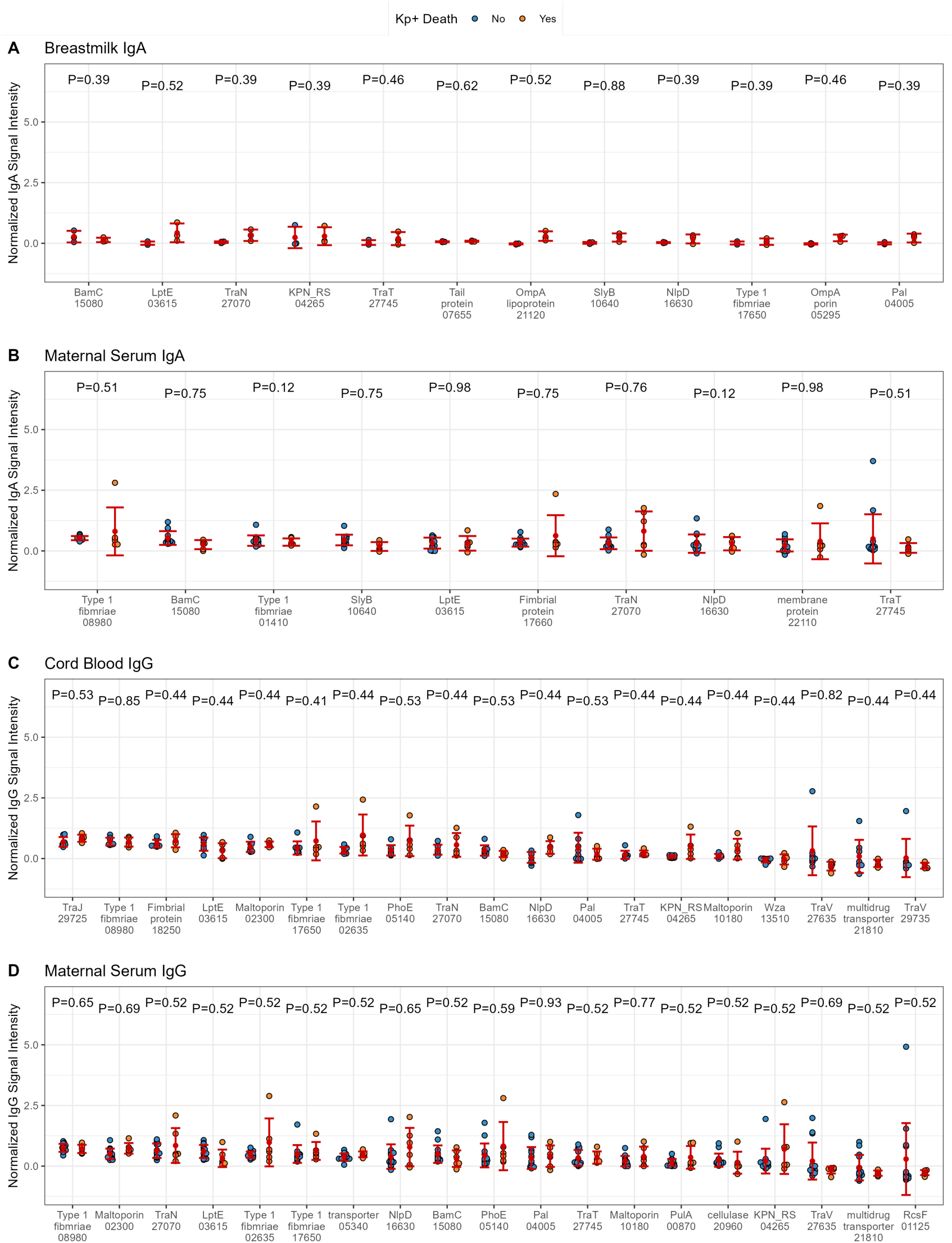
Antibody reactivity to *Kpn* proteins did not correlate with outcome of death in *Kpn*-infected infants. The beeswarm plots show the microarray signal intensity distribution of individual data points for each reactive antigen for (A) breastmilk IgA, (B) maternal serum IgA, (C) cord blood IgG and (D) maternal serum IgG. Blue points represent infants with *Kpn* sepsis that did not die, and orange points represents infants that died. Red points represent group means with error bars representing the standard deviation. Individual proteins are shown on the x-axis with protein name and RefSeq gene ID tag (5 digits following “KPN_RS”). T test P-values corrected for the false discovery rate for each comparison are shown in black above each protein.

**Figure S5.**
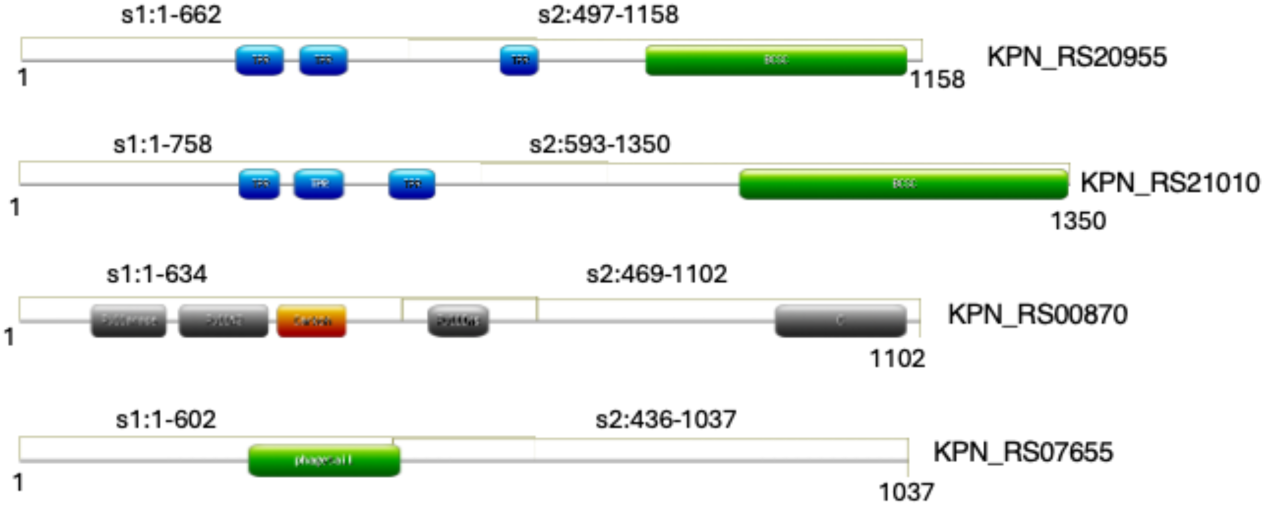
Domain layout of proteins that were also added as partial sequences. Proteins as indicated were added to the chip full-length, but also as two partial sequences to increase the likelihood of correct folding. The pearls-on-string diagram indicates the predicted domain features, the boxes overlayed visualize the two segments (s1 and s2 for each, resp.) that were also added to the chip as separate targets.

**Figure S6.**
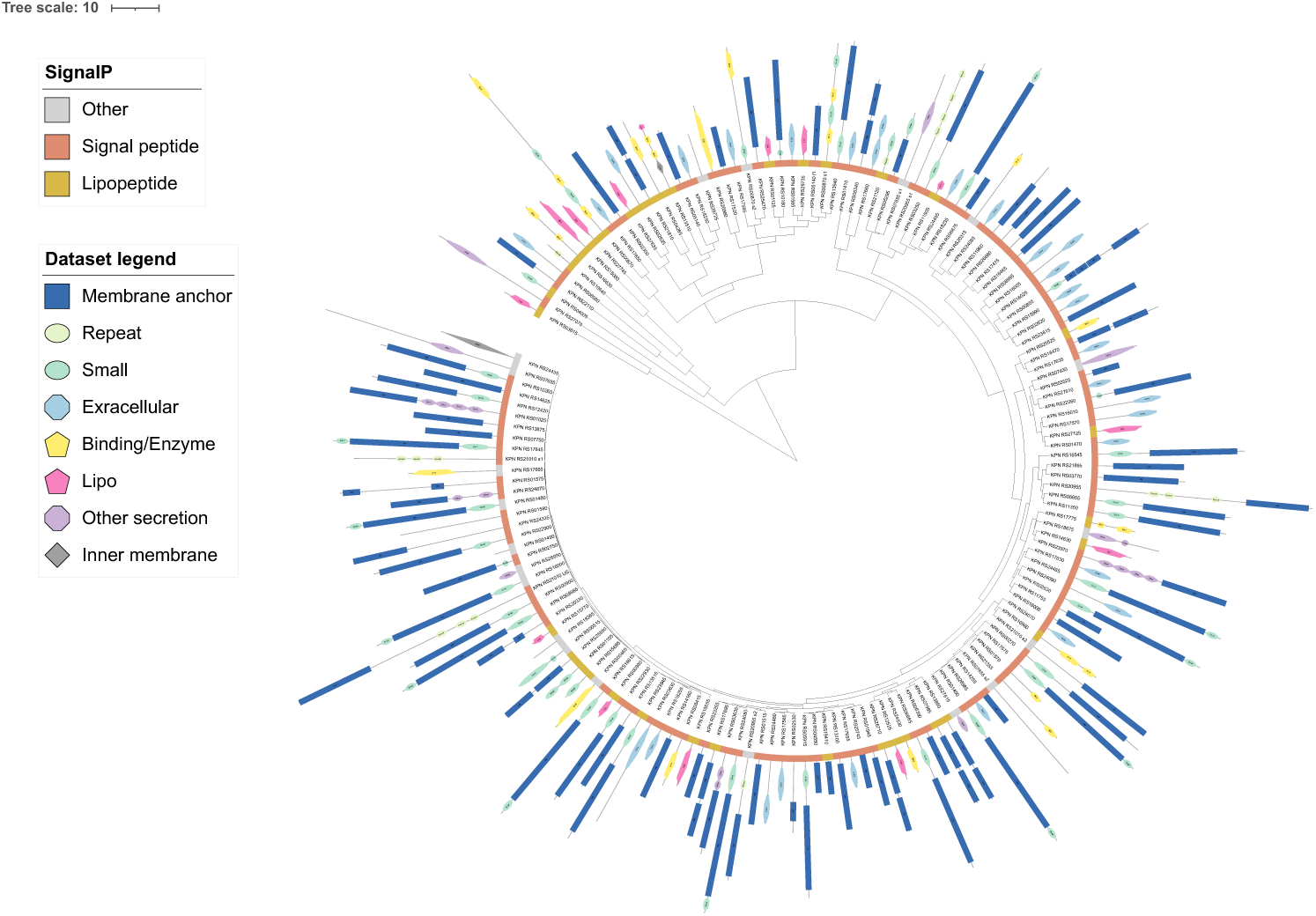
Domain architecture of all proteins on the microarray. The tree represents clustering of immune reactivity against all IgA and IgG measures combined. The domain profiles for each target are illustrated using pearl-on-string diagrams with simplified icons as indicated in the legend, representing the overall classification as in Table S2. The N-terminus of each domain architecture diagram is at the inner side of the ring, the length of the pearl-on-string icons represents the length of each sequence and domain as predicted. The colour circle visualises predicted signal sequences. The tree was visualised using iToL(1).

---

**Table S1.** Pfam domains used as search input to identify cell envelope-associated proteins.

| **Profile** | **Classification_used** |
| --- | --- |
| PF00267 | membrane_anchor |
| PF09411 | membrane_anchor |
| PF04453 | membrane_anchor |
| PF05736 | membrane_anchor |
| PF03573 | membrane_anchor |
| PF04966 | membrane_anchor |
| PF02462 | membrane_anchor |
| PF03922 | membrane_anchor |
| PF01278 | membrane_anchor |
| PF01389 | membrane_anchor |
| PF06629 | membrane_anchor |
| PF06178 | membrane_anchor |
| PF01856 | membrane_anchor |
| PF09694 | membrane_anchor |
| PF03502 | membrane_anchor |
| PF01103 | membrane_anchor |
| PF03797 | membrane_anchor |
| PF06316 | membrane_anchor |
| PF02530 | membrane_anchor |
| PF09381 | membrane_anchor |
| PF07396 | membrane_anchor |
| PF03865 | membrane_anchor |
| PF01617 | membrane_anchor |
| PF03349 | membrane_anchor |
| PF00593 | membrane_anchor |
| PF00577 | membrane_anchor |
| PF07437 | membrane_anchor |
| PF11276 | membrane_anchor |
| PF11059 | membrane_anchor |
| PF10082 | membrane_anchor |
| PF04338 | membrane_anchor |
| PF04575 | membrane_anchor |
| PF12099 | membrane_anchor |
| PF05275 | membrane_anchor |
| PF07239 | membrane_anchor |
| PF11751 | membrane_anchor |
| PF03249 | membrane_anchor |
| PF10677 | membrane_anchor |
| PF13505 | membrane_anchor |
| PF13568 | membrane_anchor |
| PF05538 | membrane_anchor |
| PF02521 | membrane_anchor |
| PF13609 | membrane_anchor |
| PF13557 | membrane_anchor |
| PF11383 | membrane_anchor |
| PF11336 | membrane_anchor |
| PF02264 | membrane_anchor |
| PF07642 | membrane_anchor |
| PF13729 | membrane_anchor |
| PF14121 | membrane_anchor |
| PF05150 | membrane_anchor |
| PF11854 | membrane_anchor |
| PF14905 | membrane_anchor |
| PF07017 | membrane_anchor |
| PF11557 | membrane_anchor |
| PF16956 | membrane_anchor |
| PF16961 | membrane_anchor |
| PF16966 | membrane_anchor |
| PF01459 | membrane_anchor |
| PF12519 | membrane_anchor |
| PF08794 | membrane_anchor |
| PF01716 | membrane_anchor |
| PF10895 | membrane_anchor |
| PF03687 | membrane_anchor |
| PF11924 | membrane_anchor |
| PF16930 | membrane_anchor |
| PF11853 | membrane_anchor |
| PF13372 | membrane_anchor |
| PF01308 | membrane_anchor |
| PF14052 | membrane_anchor |
| PF07585 | membrane_anchor |
| PF11867 | membrane_anchor |
| PF11389 | membrane_anchor |
| PF14391 | membrane_anchor |
| PF16939 | membrane_anchor |
| PF05420 | membrane_anchor |
| PF03895 | membrane_anchor |
| PF12097 | membrane_anchor |
| PF17271 | membrane_anchor |
| PF02722 | membrane_anchor |
| PF09982 | membrane_anchor |
| PF15283 | membrane_anchor |
| PF02253 | membrane_anchor |
| PF03783 | membrane_anchor |
| PF02321 | membrane_anchor |
| PF13629 | membrane_anchor |
| PF00263 | membrane_anchor |
| PF03958 | membrane_anchor |
| PF03524 | membrane_anchor |
| PF15976 | membrane_anchor |
| PF16967 | membrane_anchor |
| PF05662 | membrane_anchor |
| PF02707 | membrane_anchor |
| PF05307 | extracellular_profile |
| PF00114 | extracellular_profile |
| PF08805 | extracellular_profile |
| PF05946 | extracellular_profile |
| PF16732 | extracellular_profile |
| PF16734 | extracellular_profile |
| PF11530 | extracellular_profile |
| PF00419 | extracellular_profile |
| PF05737 | extracellular_profile |
| PF04619 | extracellular_profile |
| PF03627 | extracellular_profile |
| PF06551 | extracellular_profile |
| PF05229 | extracellular_profile |
| PF09160 | extracellular_profile |
| PF10988 | extracellular_profile |
| PF13349 | extracellular_profile |
| PF16970 | extracellular_profile |
| PF05775 | extracellular_profile |
| PF09255 | extracellular_profile |
| PF09222 | extracellular_profile |
| PF09460 | extracellular_profile |
| PF10425 | extracellular_profile |
| PF05211 | extracellular_profile |
| PF07602 | extracellular_profile |
| PF05594 | extracellular_profile |
| PF05860 | extracellular_profile |
| PF03212 | extracellular_profile |
| PF12708 | extracellular_profile |
| PF02415 | extracellular_profile |
| PF13332 | extracellular_profile |
| PF09251 | extracellular_profile |
| PF07523 | extracellular_profile |
| PF02368 | extracellular_profile |
| PF02369 | extracellular_profile |
| PF09134 | extracellular_profile |
| PF07705 | extracellular_profile |
| PF01345 | extracellular_profile |
| PF13205 | extracellular_profile |
| PF13573 | extracellular_profile |
| PF12245 | extracellular_profile |
| PF13750 | extracellular_profile |
| PF13752 | extracellular_profile |
| PF13753 | extracellular_profile |
| PF13585 | extracellular_profile |
| PF13860 | extracellular_profile |
| PF15418 | extracellular_profile |
| PF16184 | extracellular_profile |
| PF16640 | extracellular_profile |
| PF07012 | extracellular_profile |
| PF09392 | extracellular_profile |
| PF04717 | extracellular_profile |
| PF05638 | extracellular_profile |
| PF11550 | extracellular_profile |
| PF05689 | extracellular_profile |
| PF05688 | extracellular_profile |
| PF05658 | extracellular_profile |
| PF16168 | extracellular_profile |
| PF02395 | extracellular_profile |
| PF00700 | extracellular_profile |
| PF08884 | extracellular_profile |
| PF00669 | extracellular_profile |
| PF07196 | extracellular_profile |
| PF07195 | extracellular_profile |
| PF02465 | extracellular_profile |
| PF12951 | extracellular_profile |
| PF11612 | other_secretion |
| PF02501 | other_secretion |
| PF08334 | other_secretion |
| PF04956 | other_secretion |
| PF07996 | other_secretion |
| PF07244 | other_secretion |
| PF08479 | other_secretion |
| PF08478 | other_secretion |
| PF17243 | other_secretion |
| PF05947 | other_secretion |

**Table S2.** Pfam protein domains predicted for the proteins represented on the protein microarray using InterProScan.

| ID | length | Pfam profile | start | end | match |
| --- | --- | --- | --- | --- | --- |
| KPN_RS05340 | 478 | PF02321 | 282 | 452 | 5.00E-11 |
| KPN_RS05340 | 478 | PF02321 | 72 | 252 | 5.90E-24 |
| KPN_RS20955_s1 | 662 | PF13432 | 276 | 338 | 9.30E-05 |
| KPN_RS20955_s1 | 662 | PF13432 | 357 | 418 | 0.63 |
| KPN_RS20955_s1 | 662 | PF14559 | 615 | 659 | 5.70E-04 |
| KPN_RS20955_s1 | 662 | PF14559 | 473 | 532 | 0.047 |
| KPN_RS13515 | 477 | PF14052 | 89 | 467 | 3.50E-112 |
| KPN_RS01105 | 239 | PF04170 | 51 | 135 | 5.30E-23 |
| KPN_RS01105 | 239 | PF17185 | 150 | 238 | 7.10E-28 |
| KPN_RS01470 | 172 | PF00419 | 28 | 171 | 1.20E-19 |
| KPN_RS21120 | 220 | PF13488 | 37 | 85 | 4.50E-07 |
| KPN_RS21120 | 220 | PF00691 | 115 | 210 | 8.00E-21 |
| KPN_RS23255 | 286 | PF02253 | 41 | 282 | 1.60E-85 |
| KPN_RS18255 | 182 | PF00419 | 30 | 182 | 8.70E-10 |
| KPN_RS25525 | 177 | PF08212 | 35 | 174 | 3.80E-41 |
| KPN_RS13940 | 657 | PF00593 | 231 | 630 | 1.20E-59 |
| KPN_RS13940 | 657 | PF07715 | 43 | 151 | 3.90E-30 |
| KPN_RS06390 | 248 | PF06629 | 28 | 248 | 5.30E-50 |
| KPN_RS10640 | 155 | PF05433 | 62 | 102 | 3.40E-04 |
| KPN_RS26660 | 298 | PF05275 | 94 | 298 | 1.10E-83 |
| KPN_RS08980 | 334 | PF00419 | 193 | 334 | 2.70E-11 |
| KPN_RS18250 | 192 | PF00419 | 37 | 192 | 8.00E-10 |
| KPN_RS24090 | 365 | PF00419 | 204 | 365 | 1.10E-08 |
| KPN_RS23815 | 429 | PF02264 | 28 | 429 | 3.90E-129 |
| KPN_RS14625 | 440 | PF03349 | 16 | 440 | 8.10E-154 |
| KPN_RS15655 | 245 | PF13525 | 28 | 236 | 4.00E-81 |
| KPN_RS04550 | 170 | PF13505 | 9 | 170 | 2.30E-22 |
| KPN_RS20330 | 414 | PF03958 | 123 | 182 | 6.00E-05 |
| KPN_RS20330 | 414 | PF00263 | 252 | 411 | 1.30E-44 |
| KPN_RS12525 | 193 | PF03843 | 20 | 172 | 2.90E-42 |
| KPN_RS26710 | 461 | PF02321 | 274 | 456 | 6.20E-27 |
| KPN_RS26710 | 461 | PF02321 | 66 | 248 | 3.20E-17 |
| KPN_RS18235 | 864 | PF00577 | 202 | 767 | 1.50E-171 |
| KPN_RS18235 | 864 | PF13953 | 776 | 842 | 3.90E-17 |
| KPN_RS18235 | 864 | PF13954 | 37 | 182 | 6.10E-38 |
| KPN_RS17585 | 202 | PF00419 | 33 | 202 | 6.40E-06 |
| KPN_RS00465 | 313 | PF01795 | 5 | 310 | 2.10E-117 |
| KPN_RS02900 | 732 | PF00593 | 248 | 700 | 1.20E-30 |
| KPN_RS02900 | 732 | PF07715 | 83 | 180 | 1.70E-12 |
| KPN_RS26990 | 101 | PF07178 | 9 | 100 | 1.10E-28 |
| KPN_RS29735 | 189 | PF09676 | 25 | 183 | 1.80E-16 |
| KPN_RS03250 | 742 | PF00593 | 284 | 698 | 6.40E-48 |
| KPN_RS03250 | 742 | PF07715 | 54 | 168 | 2.50E-25 |
| KPN_RS05295 | 356 | PF00691 | 232 | 327 | 7.40E-21 |
| KPN_RS05295 | 356 | PF01389 | 23 | 204 | 1.30E-73 |
| KPN_RS04005 | 174 | PF00691 | 73 | 168 | 2.80E-18 |
| KPN_RS24480 | 338 | PF00419 | 184 | 336 | 1.50E-17 |
| KPN_RS08995 | 187 | PF00419 | 31 | 186 | 3.60E-17 |
| KPN_RS01480 | 827 | PF13953 | 744 | 808 | 1.80E-18 |
| KPN_RS01480 | 827 | PF00577 | 179 | 734 | 1.30E-186 |
| KPN_RS01480 | 827 | PF13954 | 18 | 163 | 1.30E-37 |
| KPN_RS07965 | 459 | PF02321 | 57 | 244 | 8.80E-18 |
| KPN_RS07965 | 459 | PF02321 | 265 | 450 | 2.10E-19 |
| KPN_RS22260 | 544 | PF11471 | 29 | 57 | 7.90E-05 |
| KPN_RS22260 | 544 | PF02264 | 127 | 544 | 2.00E-84 |
| KPN_RS00900 | 735 | PF07715 | 77 | 182 | 2.90E-22 |
| KPN_RS00900 | 735 | PF00593 | 255 | 704 | 1.90E-58 |
| KPN_RS16630 | 378 | PF01476 | 116 | 158 | 3.50E-07 |
| KPN_RS16630 | 378 | PF01551 | 278 | 371 | 6.00E-30 |
| KPN_RS03770 | 467 | PF03573 | 33 | 432 | 8.50E-21 |
| KPN_RS11350 | 717 | PF00593 | 244 | 684 | 1.30E-63 |
| KPN_RS11350 | 717 | PF07715 | 75 | 172 | 3.00E-13 |
| KPN_RS21895 | 459 | PF02264 | 82 | 459 | 2.40E-81 |
| KPN_RS20955_s2 | 662 | PF05420 | 306 | 641 | 3.60E-119 |
| KPN_RS20955_s2 | 662 | PF14559 | 119 | 169 | 1.00E-04 |
| KPN_RS00870_s1 | 634 | PF03714 | 89 | 180 | 5.40E-06 |
| KPN_RS00870_s1 | 634 | PF02922 | 316 | 400 | 2.80E-15 |
| KPN_RS00870_s1 | 634 | PF17967 | 196 | 305 | 4.00E-33 |
| KPN_RS00870_s1 | 634 | PF18494 | 501 | 574 | 9.80E-27 |
| KPN_RS24435 | 416 | PF13520 | 9 | 409 | 2.50E-25 |
| KPN_RS16000 | 845 | PF13954 | 41 | 162 | 8.30E-27 |
| KPN_RS16000 | 845 | PF13953 | 758 | 818 | 1.70E-10 |
| KPN_RS16000 | 845 | PF00577 | 177 | 749 | 3.60E-173 |
| KPN_RS23945 | 752 | PF00593 | 290 | 706 | 2.90E-49 |
| KPN_RS23945 | 752 | PF07715 | 61 | 175 | 2.10E-22 |
| KPN_RS02630 | 811 | PF00577 | 178 | 715 | 5.80E-129 |
| KPN_RS02630 | 811 | PF13954 | 26 | 161 | 6.50E-25 |
| KPN_RS02630 | 811 | PF13953 | 725 | 787 | 4.30E-15 |
| KPN_RS14255 | 180 | PF07437 | 1 | 180 | 4.30E-74 |
| KPN_RS24085 | 170 | PF00419 | 22 | 170 | 3.30E-09 |
| KPN_RS17575 | 828 | PF13954 | 37 | 181 | 3.70E-33 |
| KPN_RS17575 | 828 | PF00577 | 198 | 751 | 1.30E-190 |
| KPN_RS17575 | 828 | PF13953 | 761 | 816 | 6.80E-04 |
| KPN_RS17655 | 166 | PF00419 | 29 | 165 | 2.70E-05 |
| KPN_RS00870 | 1102 | PF11852 | 925 | 1085 | 1.10E-42 |
| KPN_RS00870 | 1102 | PF02922 | 316 | 400 | 6.00E-15 |
| KPN_RS00870 | 1102 | PF17967 | 196 | 305 | 8.80E-33 |
| KPN_RS00870 | 1102 | PF18494 | 501 | 574 | 2.00E-26 |
| KPN_RS00870 | 1102 | PF03714 | 89 | 180 | 1.20E-05 |
| KPN_RS26985 | 122 | PF05513 | 1 | 120 | 1.90E-65 |
| KPN_RS17650 | 175 | PF00419 | 27 | 175 | 1.00E-23 |
| KPN_RS15990 | 182 | PF00419 | 28 | 179 | 5.70E-16 |
| KPN_RS10180 | 538 | PF11471 | 26 | 56 | 4.70E-06 |
| KPN_RS10180 | 538 | PF02264 | 109 | 538 | 1.30E-91 |
| KPN_RS01570 | 222 | PF18649 | 150 | 221 | 2.50E-29 |
| KPN_RS11755 | 252 | PF04338 | 47 | 251 | 8.20E-54 |
| KPN_RS01125 | 135 | PF16358 | 26 | 133 | 1.70E-46 |
| KPN_RS02750 | 505 | PF02264 | 97 | 505 | 2.80E-94 |
| KPN_RS02750 | 505 | PF11471 | 28 | 58 | 5.40E-08 |
| KPN_RS25550 | 461 | PF02321 | 276 | 456 | 8.40E-24 |
| KPN_RS25550 | 461 | PF02321 | 66 | 248 | 7.80E-16 |
| KPN_RS03525 | 189 | PF07017 | 40 | 186 | 3.60E-65 |
| KPN_RS05140_f1 | 308 | PF00267 | 28 | 292 | 5.20E-99 |
| KPN_RS05140 | 308 | PF00267 | 28 | 292 | 5.20E-99 |
| KPN_RS24530 | 177 | PF08212 | 34 | 174 | 3.80E-36 |
| KPN_RS01575 | 841 | PF16967 | 283 | 350 | 7.70E-24 |
| KPN_RS01575 | 841 | PF15976 | 732 | 824 | 7.70E-28 |
| KPN_RS14630 | 253 | PF04333 | 31 | 224 | 1.70E-63 |
| KPN_RS00880 | 124 | PF09691 | 15 | 118 | 5.10E-29 |
| KPN_RS27070 | 651 | PF06986 | 355 | 600 | 1.30E-80 |
| KPN_RS17660 | 300 | PF09160 | 24 | 168 | 6.80E-65 |
| KPN_RS17660 | 300 | PF00419 | 178 | 300 | 9.00E-05 |
| KPN_RS00855 | 657 | PF03958 | 192 | 261 | 6.10E-13 |
| KPN_RS00855 | 657 | PF03958 | 126 | 188 | 1.40E-10 |
| KPN_RS00855 | 657 | PF03958 | 266 | 343 | 1.50E-13 |
| KPN_RS00855 | 657 | PF00263 | 427 | 592 | 3.20E-46 |
| KPN_RS00855 | 657 | PF21305 | 30 | 99 | 9.70E-23 |
| KPN_RS21010_US | 1350 | PF05420 | 926 | 1347 | 4.90E-110 |
| KPN_RS21010_US | 1350 | PF13432 | 354 | 418 | 2.7 |
| KPN_RS21010_US | 1350 | PF14559 | 475 | 535 | 9.80E-05 |
| KPN_RS21010_US | 1350 | PF14559 | 283 | 336 | 0.029 |
| KPN_RS16025 | 193 | PF00419 | 45 | 193 | 5.70E-18 |
| KPN_RS17665 | 470 | PF00563 | 217 | 448 | 9.70E-50 |
| KPN_RS07655 | 1037 | PF06791 | 267 | 445 | 1.60E-35 |
| KPN_RS00515 | 306 | PF01820 | 51 | 86 | 8.90E-07 |
| KPN_RS00515 | 306 | PF07478 | 103 | 302 | 3.10E-65 |
| KPN_RS17635 | 178 | PF00419 | 33 | 178 | 4.30E-26 |
| KPN_RS06655 | 727 | PF07715 | 51 | 165 | 3.40E-26 |
| KPN_RS06655 | 727 | PF00593 | 257 | 697 | 4.00E-48 |
| KPN_RS08415 | 270 | PF13365 | 66 | 237 | 0.005 |
| KPN_RS25470 | 460 | PF02264 | 83 | 460 | 2.40E-81 |
| KPN_RS11505 | 78 | PF04728 | 26 | 78 | 1.00E-22 |
| KPN_RS08985 | 833 | PF13954 | 30 | 175 | 5.00E-36 |
| KPN_RS08985 | 833 | PF00577 | 191 | 743 | 3.00E-165 |
| KPN_RS08985 | 833 | PF13953 | 753 | 813 | 3.00E-16 |
| KPN_RS16015 | 851 | PF13953 | 762 | 832 | 1.00E-12 |
| KPN_RS16015 | 851 | PF00577 | 208 | 752 | 1.10E-179 |
| KPN_RS16015 | 851 | PF13954 | 46 | 191 | 8.10E-38 |
| KPN_RS16545 | 787 | PF00593 | 277 | 744 | 2.00E-36 |
| KPN_RS16545 | 787 | PF07715 | 60 | 182 | 2.10E-07 |
| KPN_RS13100 | 381 | PF00267 | 27 | 381 | 5.10E-141 |
| KPN_RS24080 | 859 | PF00577 | 209 | 766 | 1.70E-167 |
| KPN_RS24080 | 859 | PF13953 | 776 | 843 | 9.00E-17 |
| KPN_RS24080 | 859 | PF13954 | 46 | 191 | 9.20E-35 |
| KPN_RS17565 | 211 | PF00419 | 37 | 210 | 0.0061 |
| KPN_RS18675 | 385 | PF12571 | 1 | 147 | 1.90E-41 |
| KPN_RS18675 | 385 | PF03406 | 179 | 218 | 3.30E-10 |
| KPN_RS22930 | 618 | PF00593 | 188 | 592 | 1.40E-36 |
| KPN_RS22930 | 618 | PF07715 | 40 | 146 | 7.10E-24 |
| KPN_RS16010 | 364 | PF00419 | 218 | 364 | 1.20E-12 |
| KPN_RS20955 | 1158 | PF14559 | 615 | 664 | 2.10E-04 |
| KPN_RS20955 | 1158 | PF13432 | 357 | 418 | 1.3 |
| KPN_RS20955 | 1158 | PF13432 | 276 | 338 | 1.90E-04 |
| KPN_RS20955 | 1158 | PF05420 | 802 | 1137 | 1.20E-118 |
| KPN_RS16960 | 365 | PF06725 | 277 | 354 | 6.60E-21 |
| KPN_RS16960 | 365 | PF03562 | 118 | 257 | 1.50E-32 |
| KPN_RS27745 | 243 | PF05818 | 32 | 243 | 5.20E-79 |
| KPN_RS27125 | 243 | PF05818 | 32 | 243 | 5.20E-79 |
| KPN_RS03630 | 597 | PF05860 | 28 | 275 | 1.70E-44 |
| KPN_RS16465 | 445 | PF02264 | 33 | 445 | 1.10E-87 |
| KPN_RS02530 | 381 | PF03797 | 215 | 317 | 0.019 |
| KPN_RS03615 | 196 | PF04390 | 34 | 159 | 2.00E-15 |
| KPN_RS24495 | 187 | PF00419 | 32 | 185 | 4.30E-08 |
| KPN_RS24070 | 179 | PF00419 | 30 | 178 | 2.30E-11 |
| KPN_RS01025 | 809 | PF01103 | 448 | 809 | 2.80E-106 |
| KPN_RS01025 | 809 | PF07244 | 176 | 263 | 2.10E-13 |
| KPN_RS01025 | 809 | PF07244 | 92 | 172 | 1.20E-08 |
| KPN_RS01025 | 809 | PF07244 | 266 | 344 | 2.60E-12 |
| KPN_RS01025 | 809 | PF07244 | 347 | 421 | 5.20E-11 |
| KPN_RS01515 | 350 | PF00267 | 27 | 350 | 6.10E-141 |
| KPN_RS17775 | 237 | PF01551 | 131 | 224 | 1.20E-28 |
| KPN_RS17775 | 237 | PF01476 | 30 | 72 | 7.60E-13 |
| KPN_RS06675 | 212 | PF03922 | 23 | 212 | 3.90E-72 |
| KPN_RS25745 | 181 | PF07437 | 1 | 181 | 3.20E-41 |
| KPN_RS15770 | 113 | PF04355 | 35 | 104 | 1.60E-24 |
| KPN_RS24870 | 577 | PF17243 | 26 | 100 | 3.60E-18 |
| KPN_RS24870 | 577 | PF07244 | 189 | 243 | 2.50E-07 |
| KPN_RS24870 | 577 | PF01103 | 270 | 574 | 3.00E-32 |
| KPN_RS19410 | 183 | PF07437 | 8 | 183 | 3.20E-54 |
| KPN_RS15695 | 160 | PF00691 | 55 | 151 | 7.80E-15 |
| KPN_RS06685 | 761 | PF07715 | 68 | 168 | 5.30E-18 |
| KPN_RS06685 | 761 | PF00593 | 243 | 729 | 4.00E-50 |
| KPN_RS17520 | 231 | PF06178 | 15 | 231 | 1.50E-57 |
| KPN_RS15080 | 344 | PF06804 | 41 | 342 | 2.50E-104 |
| KPN_RS02635 | 341 | PF00419 | 198 | 341 | 7.20E-15 |
| KPN_RS07630 | 310 | PF18906 | 4 | 292 | 3.40E-30 |
| KPN_RS21355 | 423 | PF01551 | 324 | 417 | 1.80E-25 |
| KPN_RS07750 | 384 | PF00267 | 27 | 384 | 1.10E-142 |
| KPN_RS24485 | 847 | PF13953 | 765 | 831 | 5.60E-17 |
| KPN_RS24485 | 847 | PF13954 | 36 | 187 | 2.50E-38 |
| KPN_RS24485 | 847 | PF00577 | 201 | 754 | 4.70E-176 |
| KPN_RS20315 | 428 | PF05036 | 344 | 418 | 6.40E-10 |
| A0A264C9S4 | 359 | PF00267 | 28 | 359 | 1.10E-122 |
| KPN_RS17570 | 331 | PF00419 | 182 | 331 | 2.70E-12 |
| KPN_RS01945 | 294 | PF03502 | 32 | 294 | 2.10E-82 |
| KPN_RS04265 | 427 | PF01095 | 193 | 341 | 5.30E-07 |
| KPN_RS05915 | 729 | PF00593 | 235 | 685 | 1.10E-29 |
| KPN_RS05915 | 729 | PF07715 | 41 | 142 | 3.60E-16 |
| KPN_RS13510 | 377 | PF10531 | 173 | 222 | 4.50E-05 |
| KPN_RS13510 | 377 | PF10531 | 256 | 310 | 41 |
| KPN_RS13510 | 377 | PF02563 | 82 | 165 | 2.10E-23 |
| KPN_RS13510 | 377 | PF18412 | 345 | 374 | 4.30E-13 |
| KPN_RS10265 | 701 | PF00593 | 231 | 663 | 4.20E-55 |
| KPN_RS10265 | 701 | PF07715 | 42 | 152 | 2.50E-18 |
| KPN_RS18470 | 490 | PF02321 | 26 | 209 | 1.80E-24 |
| KPN_RS18470 | 490 | PF02321 | 232 | 426 | 7.20E-30 |
| KPN_RS21810 | 454 | PF02321 | 270 | 442 | 1.90E-27 |
| KPN_RS21810 | 454 | PF02321 | 58 | 240 | 8.30E-16 |
| KPN_RS23970 | 807 | PF07244 | 176 | 263 | 2.70E-12 |
| KPN_RS23970 | 807 | PF07244 | 92 | 172 | 7.70E-10 |
| KPN_RS23970 | 807 | PF07244 | 347 | 421 | 1.20E-11 |
| KPN_RS23970 | 807 | PF07244 | 266 | 344 | 0.021 |
| KPN_RS23970 | 807 | PF01103 | 448 | 807 | 3.00E-94 |
| KPN_RS27635 | 194 | PF09676 | 25 | 188 | 1.30E-24 |
| KPN_RS17475 | 447 | PF16966 | 83 | 447 | 1.10E-186 |
| KPN_RS01565 | 195 | PF16449 | 26 | 193 | 1.20E-90 |
| KPN_RS01410 | 178 | PF00419 | 28 | 177 | 3.70E-14 |
| KPN_RS00270 | 782 | PF04453 | 309 | 693 | 4.50E-115 |
| KPN_RS00270 | 782 | PF03968 | 53 | 194 | 6.70E-21 |
| KPN_RS17630 | 182 | PF00419 | 33 | 182 | 1.40E-26 |
| KPN_RS16005 | 342 | PF00419 | 201 | 342 | 1.40E-06 |
| KPN_RS12420 | 706 | PF07715 | 62 | 168 | 8.90E-24 |
| KPN_RS12420 | 706 | PF00593 | 245 | 675 | 1.20E-50 |
| KPN_RS01400 | 851 | PF13953 | 770 | 833 | 3.00E-17 |
| KPN_RS01400 | 851 | PF00577 | 189 | 761 | 2.10E-143 |
| KPN_RS01400 | 851 | PF13954 | 30 | 169 | 1.40E-25 |
| KPN_RS27615 | 101 | PF07178 | 9 | 100 | 1.30E-28 |
| KPN_RS02300 | 423 | PF02264 | 33 | 423 | 1.10E-109 |
| KPN_RS17645 | 870 | PF13953 | 791 | 855 | 1.70E-19 |
| KPN_RS17645 | 870 | PF00577 | 200 | 782 | 8.40E-217 |
| KPN_RS17645 | 870 | PF13954 | 40 | 184 | 1.80E-47 |
| KPN_RS22110 | 377 | PF01103 | 137 | 377 | 1.40E-09 |
| KPN_RS17695 | 482 | PF02321 | 70 | 268 | 8.50E-16 |
| KPN_RS17695 | 482 | PF02321 | 297 | 476 | 4.30E-12 |
| KPN_RS13875 | 454 | PF04966 | 82 | 454 | 4.10E-67 |
| KPN_RS27610 | 122 | PF05513 | 1 | 120 | 1.00E-66 |
| KPN_RS11960 | 460 | PF11924 | 59 | 335 | 7.70E-102 |
| KPN_RS03635 | 556 | PF17287 | 149 | 202 | 2.50E-12 |
| KPN_RS03635 | 556 | PF03865 | 207 | 520 | 2.00E-87 |
| KPN_RS03635 | 556 | PF08479 | 74 | 146 | 3.40E-09 |
| KPN_RS13850 | 479 | PF02321 | 291 | 471 | 1.20E-10 |
| KPN_RS13850 | 479 | PF02321 | 67 | 264 | 1.90E-13 |
| KPN_RS21010_s2 | 758 | PF05420 | 334 | 755 | 1.50E-110 |
| KPN_RS24335 | 822 | PF21197 | 529 | 822 | 8.40E-129 |
| KPN_RS18000 | 422 | PF03573 | 27 | 420 | 9.40E-120 |
| KPN_RS07655_s1 | 602 | PF06791 | 267 | 445 | 6.20E-36 |
| KPN_RS14160 | 367 | PF00267 | 27 | 367 | 1.30E-149 |
| KPN_RS00870_s2 | 634 | PF11852 | 457 | 617 | 4.60E-43 |
| KPN_RS00870_s2 | 634 | PF18494 | 33 | 106 | 9.80E-27 |
| KPN_RS21010_s1 | 758 | PF14559 | 397 | 451 | 0.057 |
| KPN_RS21010_s1 | 758 | PF14559 | 283 | 336 | 0.012 |
| KPN_RS21010_s1 | 758 | PF14559 | 475 | 535 | 4.80E-05 |
| KPN_RS02620 | 186 | PF16970 | 51 | 186 | 1.10E-13 |
| KPN_RS20960 | 369 | PF01270 | 2 | 347 | 3.90E-111 |

**Table S3.** Genome sequence data accession numbers.

| Read accession | study | bioproject | biosample | sample_name |
| --- | --- | --- | --- | --- |
| **SRR37357481** | SRP679185 | PRJNA1428055 | SAMN55938643 | CNS8QI |
| **SRR37357480** | SRP679185 | PRJNA1428055 | SAMN55938644 | CNS7GP |
| **SRR37357469** | SRP679185 | PRJNA1428055 | SAMN55938645 | CNS925 |
| **SRR37357468** | SRP679185 | PRJNA1428055 | SAMN55938646 | CNS94H |
| **SRR37357467** | SRP679185 | PRJNA1428055 | SAMN55938647 | CNSANC |
| **SRR37357466** | SRP679185 | PRJNA1428055 | SAMN55938648 | CNSARR |
| **SRR37357465** | SRP679185 | PRJNA1428055 | SAMN55938649 | CNS9S3 |
| **SRR37357464** | SRP679185 | PRJNA1428055 | SAMN55938650 | CNS9AU |
| **SRR37357463** | SRP679185 | PRJNA1428055 | SAMN55938651 | CNSBIH |
| **SRR37357462** | SRP679185 | PRJNA1428055 | SAMN55938652 | CNSC45 |
| **SRR37357479** | SRP679185 | PRJNA1428055 | SAMN55938653 | BKRSSH |
| **SRR37357478** | SRP679185 | PRJNA1428055 | SAMN55938654 | CNSD5J |
| **SRR37357477** | SRP679185 | PRJNA1428055 | SAMN55938655 | CNSFKH |
| **SRR37357476** | SRP679185 | PRJNA1428055 | SAMN55938656 | CNSDL8 |
| **SRR37357475** | SRP679185 | PRJNA1428055 | SAMN55938657 | CNSIV9 |
| **SRR37357474** | SRP679185 | PRJNA1428055 | SAMN55938658 | CNSKU5 |
| **SRR37357473** | SRP679185 | PRJNA1428055 | SAMN55938659 | CNSJXZ |
| **SRR37357472** | SRP679185 | PRJNA1428055 | SAMN55938660 | CNSJXL |
| **SRR37357471** | SRP679185 | PRJNA1428055 | SAMN55938661 | CNSMKB |
| **SRR37357470** | SRP679185 | PRJNA1428055 | SAMN55938662 | BKRS5B |

